# Modeling Population Vulnerabilities to Climate Change-Driven Hurricanes and Tropical Storms in the North Atlantic Basin

**DOI:** 10.64898/2026.08.16.26360517

**Authors:** Shaelyn Mullins, Johnny Uelmen

## Abstract

Tropical cyclones are among the deadliest and costliest natural disasters in the United States, and the most intense storms are expected to become more frequent as the climate warms. Anticipating where deaths are most likely to occur is therefore central to preparedness, evacuation planning, and public health response. We modeled block-level mortality risk for twenty-four of the deadliest and costliest tropical cyclones to strike the U.S. Gulf and East Coasts, Puerto Rico, and the U.S. Virgin Islands between 1992 and 2024. For each storm, we combined NOAA hazard data (wind swaths, rainfall, and storm-surge inundation) with 2020 U.S. Census demographic and socioeconomic characteristics and the CDC/ATSDR Social Vulnerability Index for all Census blocks within 25 miles of the coast, and trained storm-specific boosted-tree models with population-standardized mortality as the outcome. Averaging block-level predictions within Saffir-Simpson categories yielded risk maps spanning tropical storms through Category 5 hurricanes. Predicted mortality risk rose with storm severity and concentrated in urban coastal communities of Puerto Rico, Louisiana, Florida, North Carolina, Virginia, Maryland, New Jersey, and New York, as well as in low-lying inlet, peninsula, and sound geographies. Large block population, non-Hispanic composition, male-dominated blocks, predominantly white blocks, and males aged 20 to 34 years ranked among the strongest predictors of mortality — patterns that likely reflect structural factors shaping exposure rather than individual susceptibility. The category-specific risk maps and an accompanying interactive dashboard provide a practical decision-support tool for emergency managers, planners, and coastal residents preparing for future storms.

**Plain Language Summary:** Hurricanes and tropical storms are among the deadliest and costliest disasters in the United States, and climate change is expected to make the strongest storms more frequent. Knowing which communities are most likely to suffer deaths during a storm helps officials plan evacuations, strengthen buildings, and direct help where it is needed most. We combined records from twenty-four of the most damaging storms to strike the United States, Puerto Rico, and the U.S. Virgin Islands since 1992 with detailed information about the people living within 25 miles of the coast. For each storm, we measured rainfall, wind, and flooding, described the makeup of every neighborhood, and used a computer-learning method to estimate how at risk each small area would be. We then grouped storms by strength to map where danger is greatest for each storm category. Risk was highest in densely populated coastal cities and in low-lying inlets, peninsulas, and sounds, and it grew as storms strengthened. These maps are available in a public, interactive online dashboard so that residents, planners, and emergency managers can explore local risk and prepare before the next storm.

**Key Points:**

- Boosted-tree models estimate fine-scale mortality risk from tropical cyclones along the United States Atlantic and Gulf coasts
- Risk concentrates in urban coastal communities of Puerto Rico, Louisiana, Florida, and the Mid-Atlantic and rises with storm severity
- Category-specific risk maps and an interactive dashboard can guide preparedness, evacuation planning, and public health communication

## 1. Introduction

Severe tropical cyclones are intensifying in a warming climate: rising greenhouse gas concentrations trap outgoing longwave radiation, warming the lower atmosphere and the upper ocean and driving sea-surface temperatures further from their historical norms. Oceanic warming, exacerbated by anthropogenic activities, can influence the abiotic conditions that trigger natural disasters, particularly tropical cyclones (Franco et al., 2020). Simulations based on historical sea-surface temperature trends indicate these temperatures will continue rising as the climate warms (Alexander et al., 2020), and analyses of the North Atlantic Oscillation and multidecadal climate models support the expectation of more severe tropical storms in the future (Delworth et al., 2016). There is thus an urgent need for forecasting tools that identify the populations and environments at highest risk of adverse outcomes. Building on these works, this study examines some of the most destructive storms in recent U.S. history to better understand population vulnerability.

Mortality is the most consequential measure of tropical cyclone impact on public health. Direct mortality from tropical storms and hurricanes refers to deaths caused by flooding, drownings, rough seas, rip currents, collapsing structures, and lightning strikes during landfall (Beven et al., 2019). Indirect deaths stem from infectious diseases spread through close contact and new flooding, economic and displacement turmoil, and the exacerbation of untreated chronic illness when access to healthcare is lost; electrocutions, vehicle accidents on flooded roads, house fires, and some tornado-related deaths are also generally counted as indirect mortality (Beven et al., 2019). A Hurricane Ike case study reported as many as five indirect deaths for every one death directly attributable to the storm (Zane et al., 2011). This research examines which communities are disproportionately vulnerable to mortality during severe tropical cyclones, and these insights will guide an educational tool tailored to strengthen preparedness and community resilience.

With increasing probabilities of severe storm formation in the coming years, the potential impacts of these natural disasters could devastate lives and communities across all aspects of society. A 2017 study examined the effects of natural disasters on mental health and population displacement, finding that individuals forced to evacuate and relocate face higher risks of stress and PTSD (Schwartz et al., 2017). Additionally, a case study of Hurricane Maria, which devastated Puerto Rico in 2017, highlighted a migration crisis: over 90,000 people were displaced by storm damage, resulting in a population influx into Florida (Robins, 2024). Hori and Schafer’s 2009 report on the costs associated with the 2005 Hurricanes, Katrina and Rita, identified numerous disadvantages that encumbered communities afterward. Follow-up surveys in Louisiana revealed increased rates of unemployment and mental health issues (Hori & Schafer, 2009). These studies illustrate how tropical storms can substantially impair quality of life, affecting physical, mental, and emotional well-being and exacerbating economic hardships. This study aims to identify vulnerable communities to aid strategies that can reduce future mortality and decline in well-being, the primary outcomes of this research.

The need to sample severe storms is also motivated by damage to land cover, vegetation, biodiversity, and other living organisms from increasingly frequent and intense hurricanes. Beachy shore environments are particularly vulnerable topography that cannot be easily drained (Zhang et al., 2020) — a common characteristic of the U.S. Gulf and East Coasts and U.S. territories that heightens population vulnerability to the storms that repeatedly strike this region. We therefore sampled extreme storms affecting these geographies to understand population-level vulnerability and inform preparation and outreach that improve community-level health outcomes.

In addition to environmental damage, intense tropical cyclones can contribute to the spread of infectious diseases and epidemics. The National Institutes of Health (NIH) reported that Norovirus, Salmonella, toxigenic and nontoxigenic *V. cholerae*, and other diseases such as Hepatitis A and E, and leptospirosis (Liang and Messenger, 2018), spread throughout populations following Hurricanes Katrina (2005) and Allison (2001). Additionally, diseases with high viral loads and other respiratory infections become highly communicable as a result of new overcrowding conditions brought on by displacement due to storms (Watson et al., 2007), suggesting the need for supplemental tools to expand public health interventions for increasingly extreme events.

The economic toll of these storms, adjusted for 2023 inflation, amounts to trillions of dollars in losses for the United States (National Centers for Environmental Information, 2025). Natural disasters drive declines in gross domestic product (GDP), supply-chain disruptions, loss of vital infrastructure, agricultural damage, and setbacks to economic enterprises (Frame et al., 2020); the burden of Hurricane Harvey in 2017 alone has been estimated at 375,000 life-years lost (Frame et al., 2020). Increased urbanization along U.S. coastlines has created more opportunities for disasters to damage critical infrastructure. Higher-quality buildings generally withstand serious damage better than lower-quality housing, which is often associated with lower socioeconomic status (Salim et al., 2024), leaving these populations especially vulnerable to hurricanes. These economic burdens reduce quality of life and often compound other disadvantages, highlighting the need for tailored public health interventions and tools that model worst-case storm scenarios to aid emergency preparedness.

These mortality and economic tolls converge on healthcare systems in the aftermath of catastrophic events: as the number of people needing health services surges, access to those services simultaneously deteriorates, and loss of life grows (Shultz et al., 2018). A case study of the 2004 hurricane season in Florida attributed approximately 53% of deaths to untreated chronic conditions following the loss of healthcare infrastructure (McKinney et al., 2011). Other studies documented post-hurricane spikes in suicide (Zane et al., 2011), hospitalizations (Dosa et al., 2010), delayed cancer diagnoses and exacerbated chronic disease (Cowan et al., 2025), and heart-related deaths (McKinney et al., 2011). These increases indicate that severe storms affect health long after landfall, well beyond drownings and other deaths caused directly by peak storm conditions.

These findings underscore the need to address preventable deaths from tropical cyclones. This study identifies the Census blocks most vulnerable to storm-related mortality to support earlier intervention strategies and emergency preparedness. We model mortality as the primary outcome, using hurricane severity characteristics together with demographic and socioeconomic covariates to assess population-level vulnerability. Key hurricane features — wind swath width, mean rainfall, and storm-surge inundation — are analyzed alongside the socioeconomic makeup of populations along the Gulf and East Coasts and in Puerto Rico and the U.S. Virgin Islands, and economic burden is modeled using county GDP trends from the years preceding each storm through the storm year. Combining these elements makes it possible to estimate which populations have borne the highest costs from severe storms. With this information, we propose a public-facing tool that supplements existing forecasting strategies and incorporates community- based needs to enhance preparedness for extreme tropical cyclones. We predicted that populations with the greatest social vulnerabilities would face the highest mortality risk, that risk would increase with storm severity, and that blocks with higher Social Vulnerability Index (SVI) rankings, lower incomes, and predominantly non-white populations would be at highest risk. Analyses were performed in ArcGIS Pro (ESRI 2026), JMP (Student Edition 18.2.1), and R (RStudio 2025.08.0-357).

## 2. Methods

All data collected for this study are derived from publicly available sources accessed through the U.S. Census Bureau and the National Oceanic and Atmospheric Administration (NOAA). The finest resolution available, block level, was gathered to obtain the most accurate data on the population-level makeup of the Gulf and East Coasts, and U.S. territories. Our sampled geographies are within 25 miles of the contiguous U.S. coast and territories. We assumed that populations residing in these areas would be most susceptible to storm-related damage from wind, precipitation, storm surge, and flooding/inundation (Nam et al., 2026).

### 2.1 NOAA Data Collection

To examine which populations are particularly susceptible to severe storms, we selected twenty-four of the deadliest and costliest storms between 1992 and 2024, drawing the top five storms within each category of the Saffir-Simpson scale from NOAA’s memorandum of “Costliest U.S. Tropical Cyclones.” The Saffir–Simpson Hurricane Wind Scale ranks hurricanes in five categories (1–5) by maximum sustained wind speed; we added a tropical-storm (TS) tier below Category 1 to capture sub-hurricane systems that nonetheless caused substantial mortality and damage, yielding six severity strata. Very few Category 5 storms have made landfall on record, so only two could be sampled within the study period; similarly, only three tropical storms met the mortality and economic thresholds defined in NOAA’s memorandum (National Centers for Environmental Information, 2025). See Tables 2A and 1B for further details. Specific hurricane features were compiled from publicly available summaries in NOAA’s National Hurricane Center and Central Pacific Hurricane Center division (NHC).

The summary files provided the landfall date as the primary date extracted from the multi-day span most hurricanes encompass. Pressure and wind speed were also compiled, which contribute to the physical characteristics of hurricanes and tropical storms. Coordinates of landfall were recorded for each storm as these locations tend to bear the highest levels of destruction. Table 2A summarizes the aforementioned characteristics.

Mortality, our primary outcome, is detailed in separate files for each sampled hurricane. These data are sourced from the National Weather Service’s (NWS) Storm Events Database, which compiles weather-related deaths, is maintained by the National Centers for Environmental Information, and is verified by the CDC. Mortality is reported at different resolutions across our sample (county versus state). Because indirect mortality is likely underreported during the prolonged aftermath of highly destructive storms (such as Katrina in 2005 and Maria in 2017), we consolidated indirect deaths with direct deaths. Deaths outside the 25-mile buffer, and deaths outside the wind swath area lacking sufficient location detail, were excluded from the analysis; Table 3A summarizes the death totals retained. We therefore acknowledge that our estimates are likely conservative and that true excess mortality risk is greater than this study concludes.

Mortality was then manually assigned to the respective counties or states in the individual storm datasets in JMP and population-standardized by multiplying the unadjusted death total by the block’s population divided by the total population of the reporting state or county. This adjusted mortality was merged back into the master dataset.

Storm surge data was collated by category according to the Saffir-Simpson scale. This data represents the maximum predicted flooding and inundation that could occur based on storm severity. The NHC at NOAA generates risk maps that predict which communities are particularly vulnerable to inundation and flooding along the U.S. Gulf and East Coasts, as well as in the U.S. Virgin Islands and Puerto Rico. These digitized maps contain pixels assigned values that estimate the amount of flooding, measured in foot increments. These data were brought into R Studio to be processed and extracted at the block level. Once this was complete, the datasets were uploaded into JMP, and the following calculation was completed to estimate the area of the block affected by hypothetical inundation:

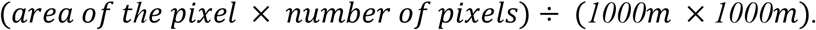

Rainfall data were collected to estimate mortality risk and contextualize sociodemographic associations, but were inconsistent across storms. Most NOAA summary files included a choropleth map of rainfall, measured in inches. Because these maps were not digitized, ArcGIS Pro was used to classify and rasterize the images: TIFF files were georeferenced onto the correct coordinates within the U.S. and its territories, the Training Samples Manager tool was used to assign rainfall values to pixels, and the Classification Wizard tool performed pixel-based classification using machine learning techniques. We then cleaned borders, track lines, and other misclassified pixels to assemble the final raster, smoothing as needed by adjusting the map symbology. Rainfall maps for pre-2009 storms were not provided in the NOAA summary files and were sourced from other NOAA archives, such as the NHC Data Archive, then georeferenced and classified in the same manner. To link rainfall to blocks within the 25-mile range, each raster was loaded into RStudio, overlaid with the Census shapefile, and the mean rainfall across the pixels making up each block was extracted. This process was repeated for each storm.

Wind swath is the final storm characteristic essential to determining storm breadth and severity. Wind swaths categorize areas by wind-speed thresholds, with higher winds posing greater risk to affected communities. For storms occurring in 2009 or later, NOAA provides wind swaths in shapefile format compatible with ArcGIS Pro; these were downloaded from NOAA’s NHC archives and loaded into ArcGIS Pro, the census buffer files were added, and blocks overlapping the swaths were extracted with the Pairwise Intersect tool and exported as a separate file aggregating block-specific and wind swath data. Swath data were unavailable for storms prior to 2009; further archival searches recovered these swaths as TIFF files, which were georeferenced in ArcGIS Pro as with the rainfall files. Shapefiles were then created using the Create Polygon feature, with inner polygons assigned “Hurricane Force Winds” (corresponding to 50 knots or more) and outer polygons “Tropical Storm Force Winds” (up to 50 knots). The 25- mile buffer layer was added to the map with the new polygons, and affected blocks were extracted using the Pairwise Intersect tool. All wind swath data were standardized to either “Tropical Storm Force Winds” or “Hurricane Force Winds” to allow meaningful comparisons between storms.

Within each summary file, the unadjusted economic cost is available. NOAA’s memorandum has economic measures in USD adjusted for 2024 inflation using the 2024 Consumer Price Index. These details are summarized in Table 2A. To further clarify the economic impact, storm-to-storm county-level GDP was obtained from the U.S. Bureau of Labor Statistics for 2001 through 2023. To estimate the effect of the storm’s impact year to year, the GDP of the year(s) prior to the storm and the year of the storm’s landfall were utilized to understand the magnitude of the impact of these tropical cyclones. Storms prior to 2001 could not include economic impacts in their models due to the lack of data.

All rainfall and wind files were processed in separate ArcGIS Pro maps, and extractions were performed within each file. The exported files were merged into a master dataset in JMP by linking on each block’s unique GeoID, generated by the U.S. Census Bureau. Population-standardized mortality was merged in the same manner. Economic data, compiled at the county level, were likewise merged by GeoID, with county values applied to all blocks within each county.

### 2.2 U.S. Census Data Collection

The U.S. Census Bureau gathers population data from each household (and group facilities), supplemented by in-person follow-up to produce a decennial report on the population makeup. The demographics collected for this study include: age, sex, ethnicity, race, household income, household ownership, and household size. Literature highlights these characteristics as essential risk factors for overall population vulnerability, particularly in the context of catastrophic natural disasters (Cowan et al., 2025). Furthermore, these risk factors are utilized in the 2022 Social Vulnerability Index (CDC, 2024), and older, lower-income, non-white populations demonstrate greater general susceptibility in terms of mortality, morbidity, and economic strain as compared to other demographics. In the U.S. Census database, we searched each demographic, applying the following filters: block-level resolution and the decennial year 2020. Block level is the finest resolution available from the U.S. Census, two orders of magnitude lower than census-tract resolution (see Fig. 1). Any state more than 25 miles inland of the Gulf and the East Coast was filtered out of the collection, as literature demonstrates this geography as historically susceptible to tropical cyclones. This data is derived from the official 2020 census. While our hurricane data begins in 1992, 2020 was used to extrapolate characteristics, as there are numerous inconsistencies across the decennial data 1990, 2000, 2010, and 2020, including conflicting spatial resolutions, stratifications, and categorizations. The 2020 data is the most comprehensive and consistent of these four censuses. Additionally, as we intend to predict vulnerabilities to future severe storms, the 2020 population makeup would more accurately reflect the population makeup in the future.

**Figure 1.**
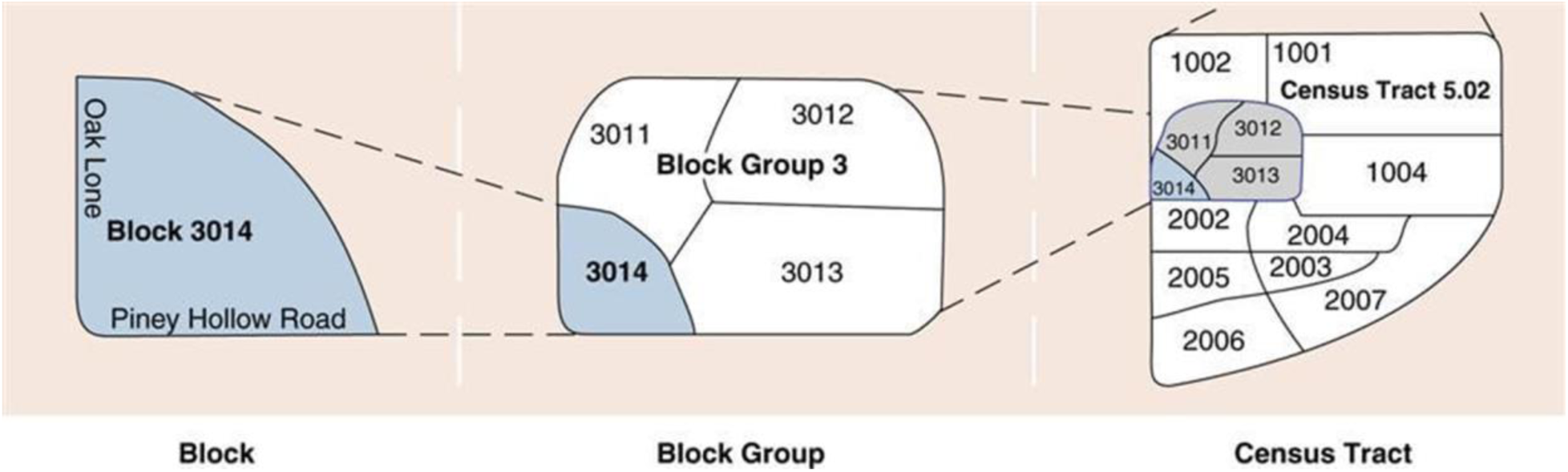
U.S. Census geographic hierarchy showing blocks (analytic unit), block groups, and Census tracts, which together form counties or equivalent jurisdictions within each state.

Age and sex were collected as a single dataset from the 2020 Census Demographic and Housing Characteristics file, where age ranges (ages 5 to 85 and 85 and over) are stratified by sex (male or female). Values represent the number of individuals in that block that fall within a given category. For example, if a given block has a value of 10 for “females aged under 5 years,” that block contains 10 females under 5 years of age. This is the same reporting mechanism for all demographics captured. Ages were collapsed into epidemiological age groups according to the National Institutes of Health (Geifman et al., 2013). Collapsed categories are as follows, measured in years: under 5, 5-14, 15-19, 20-34, 35-49, 50-64, 65-79, 80 and older. Filtered at the block level, only states within our 25-mile buffer, as well as Puerto Rico and the U.S. Virgin Islands, were selected and downloaded, then only blocks up to 25 miles inland were extracted.

Ethnicity data is characterized as “Hispanic/Latino” or “Not Hispanic/Latino” from the same 2020 Demographic and Housing Characteristics survey. Races include White, Black or African American, American Indian or Alaskan Native, Asian, Native Hawaiian and Other Pacific Islander, and Other Races.

Three key socioeconomic characteristics were collected to model structural and economic vulnerabilities within the populations of interest. Household income data from 2020 were available only at the block group level and compiled from the American Community Survey, conducted by the U.S. Census Bureau. The data covers the 12 months before the 2020 Census and, at the time of dissemination, was adjusted for 2023 inflation. Income categories ranged from less than $10,000 annually to $200,000 or more, yielding 16 income levels; these were further grouped into quartiles for easier analysis using the midpoints of the income ranges predefined by the U.S. Census Bureau. This produced 16 income levels, which were further grouped into quartiles for easier analysis. Household tenure was sampled from the 2020 Census Demographic and Housing Characteristics dataset, using the most complete data available on tenure (owner or renter), stratified by age (5 to 85 and older). Intervals varied from 2 to 5 years. This data was available at the block level and filtered using the same methodology as all other demographic data collected. Totals for owning and renting were also available at the block level and used in our dataset; other stratifications were excluded. Lastly, household size was collected at the block level for locations within a 25-mile buffer from the 2020 Decennial data file, available as household size (ranging from 1-person households to 7 or more people) stratified by tenure (renting or owning).

Once all demographic data were cleaned and filtered for the appropriate blocks, all files were merged into a single master dataset in JMP Student Edition 18.2.1.

### 2.3 Social Vulnerability Index

The 2022 Social Vulnerability Index was downloaded from the Centers for Disease Control and Prevention (CDC), and the appropriate blocks were extracted from the file to aid in identifying high-risk populations. The summary variable of interest, isolated for analysis, is an overall percentile ranking relative to all other blocks within the state. This summarizes poverty status, employment status, insurance status, age, disability status, household makeup, English language proficiency, lack of a high school diploma, housing type, transportation access, race, and ethnicity. For all these markers, a value between 0 and 1 is assigned, with values closer to 1 indicating greater vulnerability and those closer to 0 indicating lower vulnerability (CDC, 2024).

In total, the final dataset included socioeconomic and demographic variables — age, gender, race, ethnicity, household size, household income, tenure status, and SVI score — reflecting the vulnerabilities of specific blocks, alongside environmental covariates related to storm severity: wind swath width and strength, mean rainfall, and mean predicted inundation. These are summarized by SVI quintile in Table 1A, with a state-level breakdown in Table 4B. Together, these factors identify the characteristics of blocks at higher risk of excess mortality and economic burden from storms, highlighting coastal communities with greater vulnerability to future extreme storms.

### 2.4 Data Analysis

Boosted trees (JMP Student Edition 18.2.1) were fit for each storm in our sample with at least one storm-attributable death. All covariates were input with adjusted mortality as the outcome for each storm. Using 23 storm-specific boosted trees, we estimated block-level risk, capturing how vulnerable each community would be to that storm given its population profile and the mortality observed when the historical storm struck.

Boosted trees were used for their strong predictive performance and their iterative training via hundreds of decision trees. Because boosted tree models are not based on coefficient estimation, multicollinearity was not treated as a model assumption requiring exclusion of correlated predictors; however, correlated covariates were reviewed to assess redundancy and support interpretation. Covariates were excluded from the individual boosted trees if there was insufficient data for a given predictor (i.e., Hurricane Andrew did not have an economic impact variable due to a lack of data availability for 1992 from the U.S. Bureau of Labor Statistics).

Once each storm-specific risk prediction was complete, predictions were averaged within the six severity categories to estimate a block’s expected risk by storm category. For example, averaging the risk across all Category 4 storms in our sample estimates the block-level risk if a Category 4 storm, like Hurricane Maria, were to strike any other area in the 25-mile buffer (Fig. 2E). Future analyses will examine storm-specific effects and predictors.

**Figure 2.**
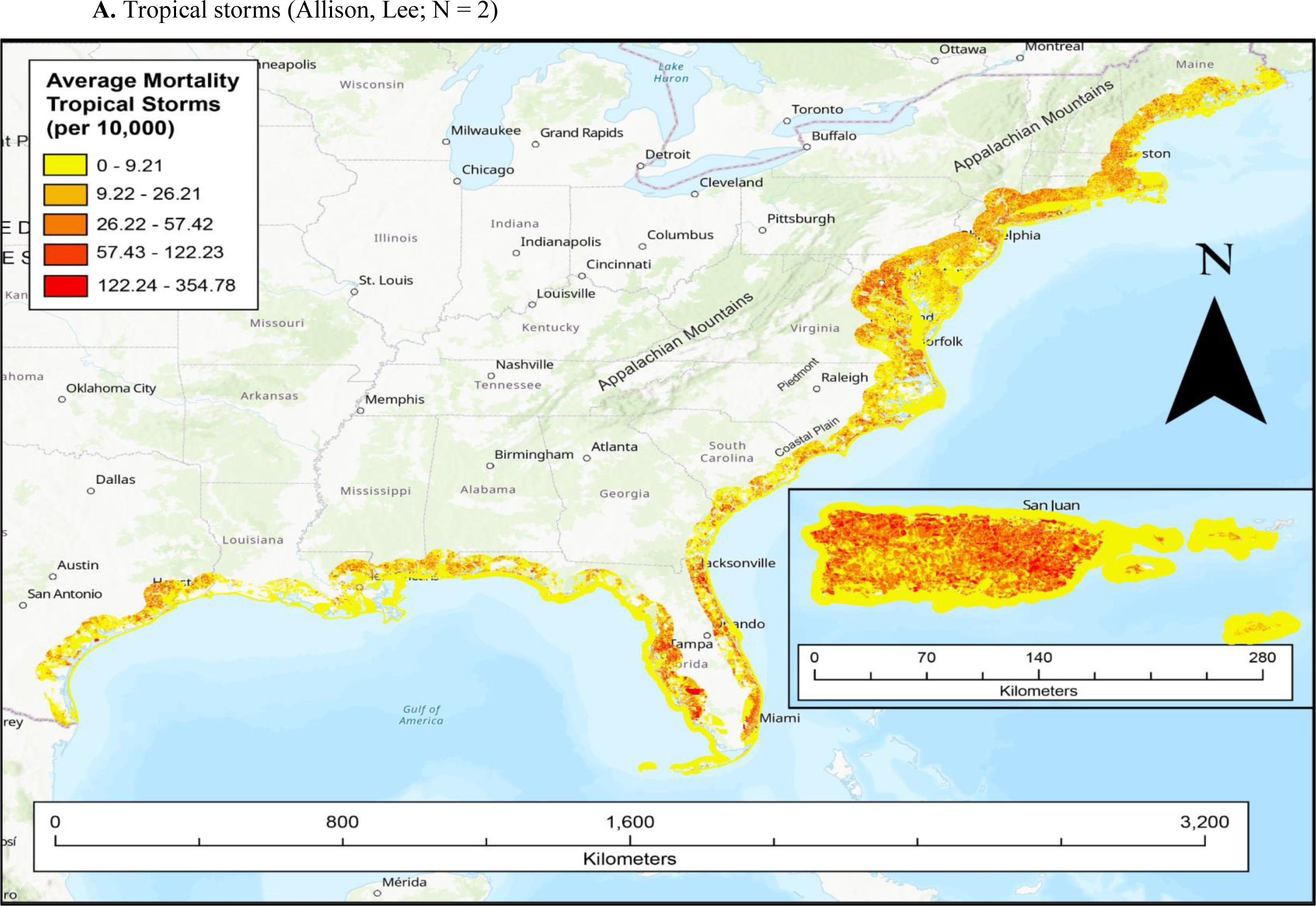
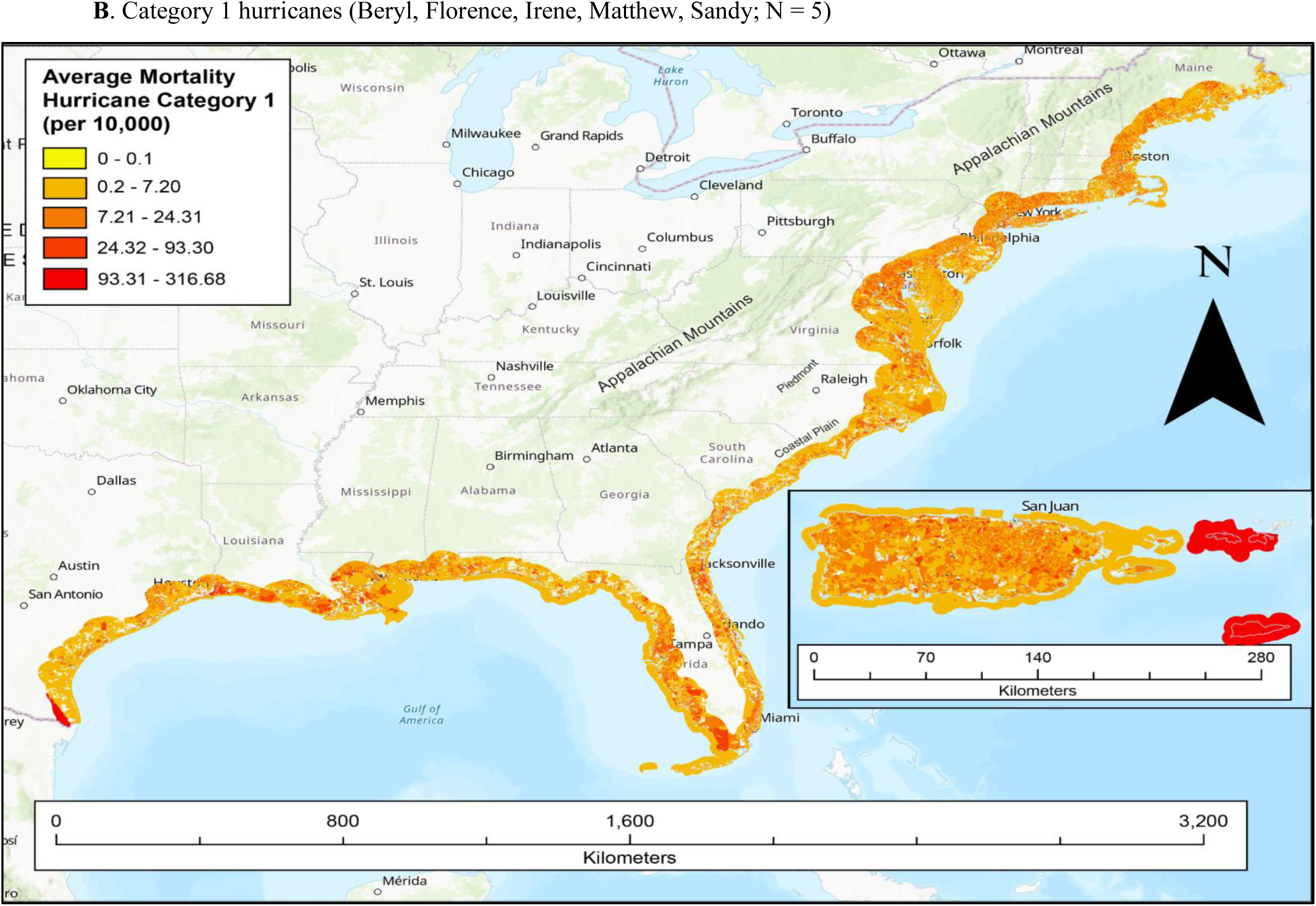
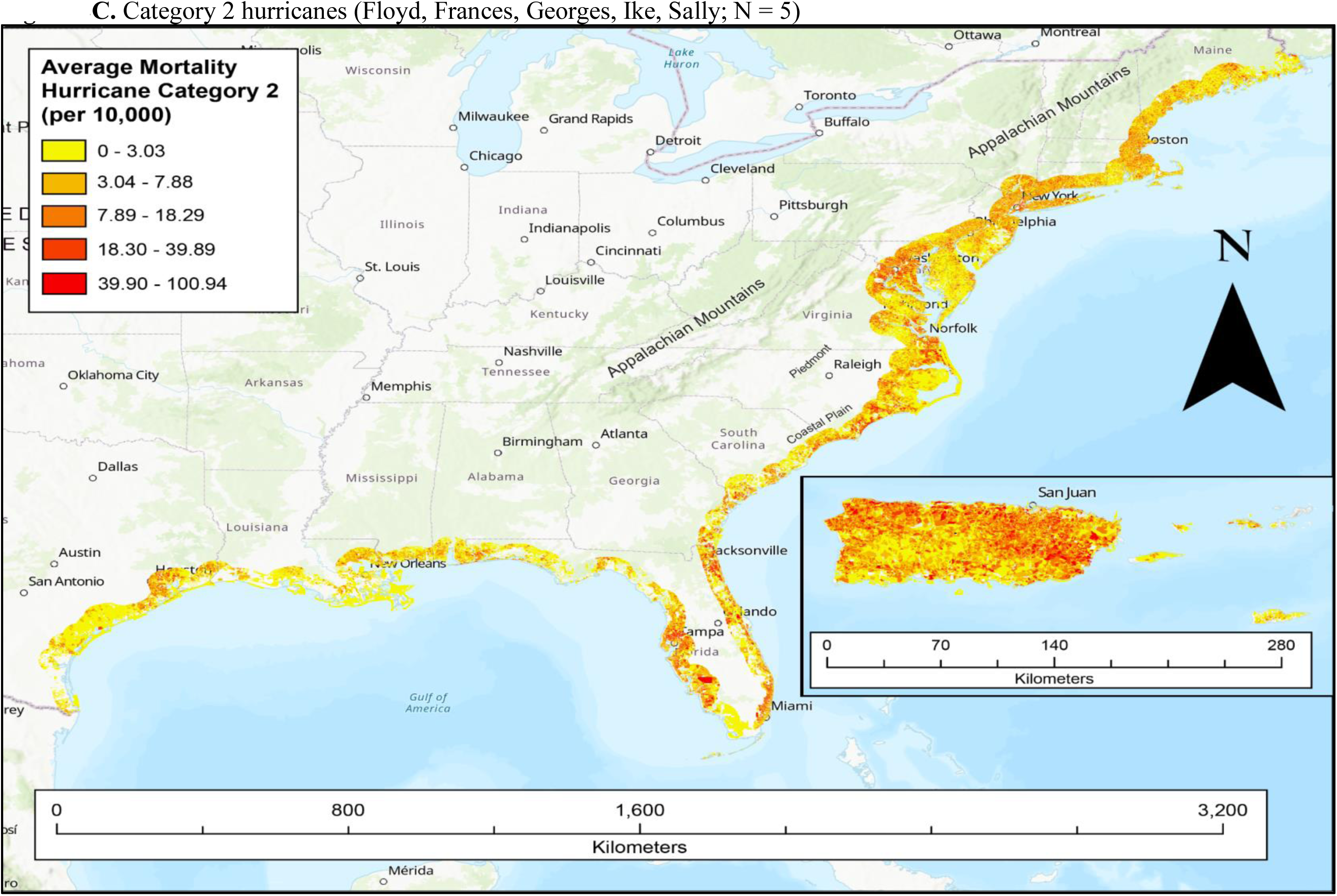
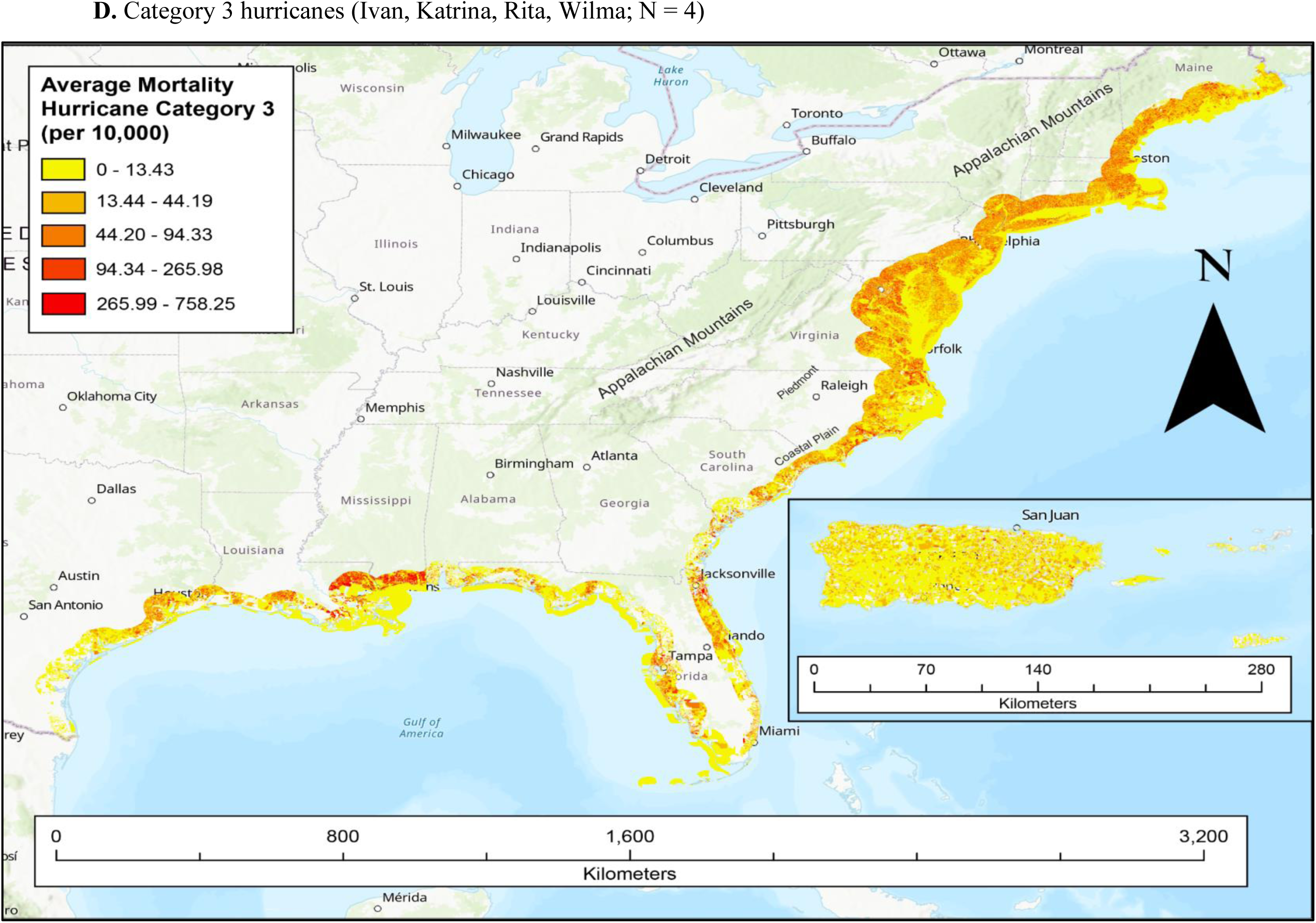
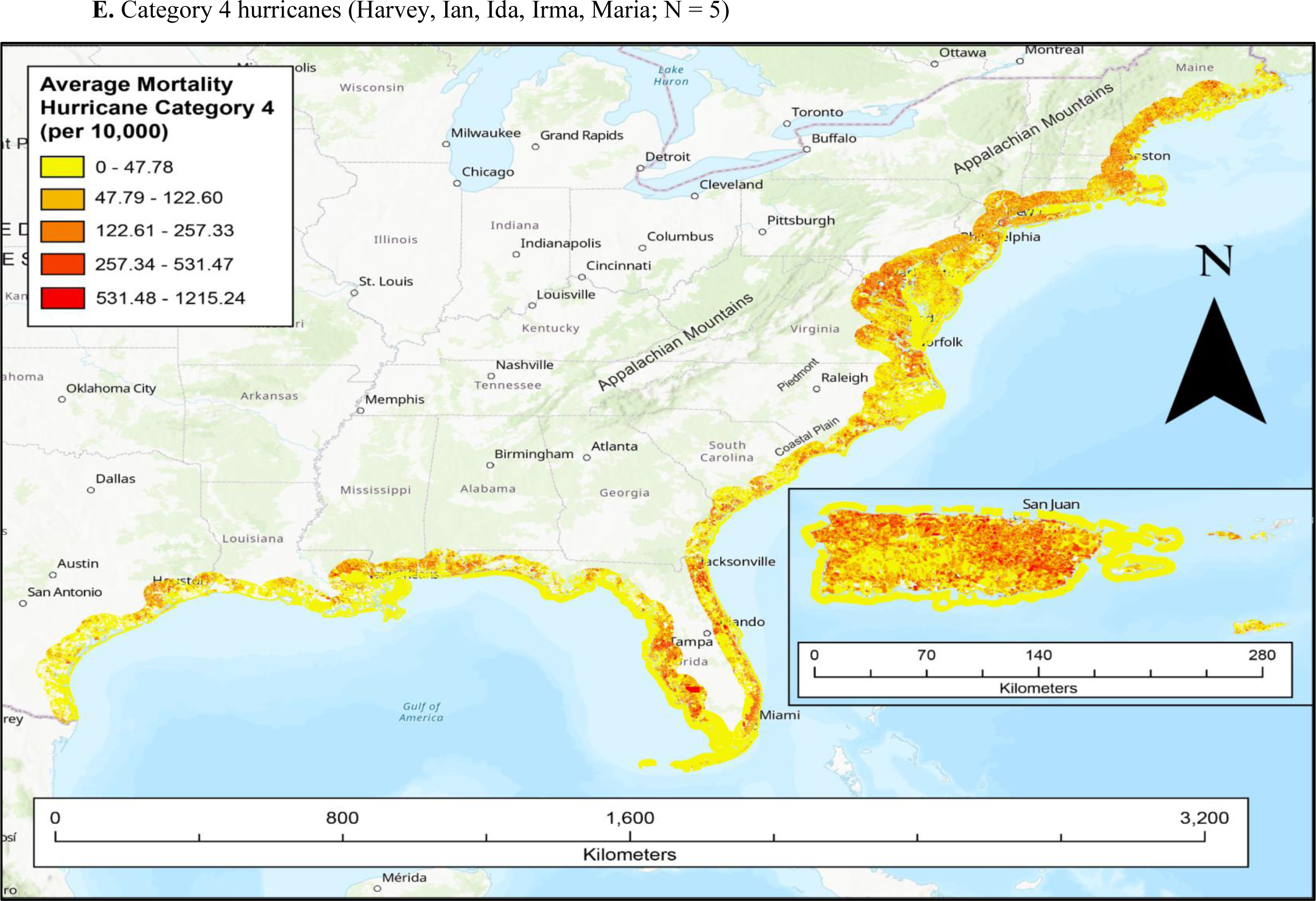
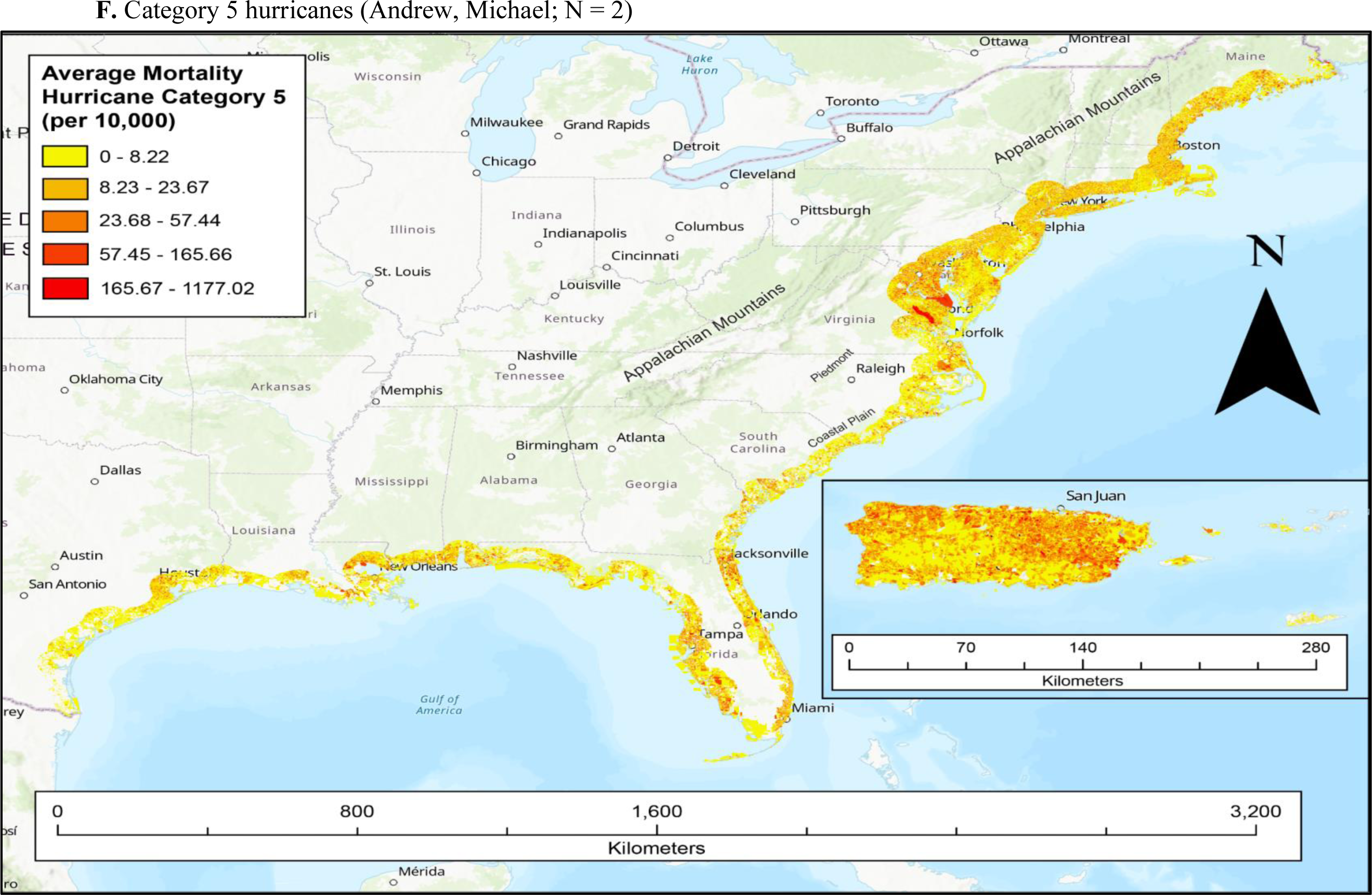
Block-level predicted mortality risk by storm category along the U.S. Gulf and East Coasts, Puerto Rico, and the U.S. Virgin Islands, within 25 miles of the coast. Panels show average predicted risk for (A) tropical storms, (B) Category 1, (C) Category 2, (D) Category 3, (E) Category 4, and (F) Category 5 hurricanes, averaged across the storms sampled in each category (N = 23 storm models; panel headings list the contributing storms). The darkest red denotes the highest predicted mortality risk and yellow the lowest. Each surface is generated from the data input to that category’s storm-specific boosted-tree models. An interactive, queryable version of these surfaces is available through the dashboard described in Section 5.

## 3. Results

Storm-specific results from twenty-three separate boosted trees are reported in Table 4A, including R^2^ and the root average square error (RASE) for each storm. Tropical Storm Imelda was excluded as no mortalities were reported within our 25-mile buffer. Across all boosted trees, RASE outputs were small, suggesting relatively accurate predictions. R^2^ varied, but overall demonstrated adequate model fitness across storms. Comparing the training and validation outputs for each storm, RASE and R^2^ values were relatively consistent, suggesting sufficient predictive accuracy. See Table 4A for specific outputs. Table 3B presents the specific variables ranked most important for predicting mortality based on the decision trees generated for each boosted tree model. This explains the relative influence of these variables in predicting mortality.

Some storms could not include all covariates due to data limitations or inconsistencies. For example, GDP could not be included as a covariate for storms occurring before 2001, because the U.S. Bureau of Labor Statistics provided data only from 2001 to 2023.

The following maps (Fig. 2A-2F) depict average predicted risk at the block level along the U.S. Gulf and East Coasts, Puerto Rico, and the U.S. Virgin Islands, by storm category. An interactive, queryable version of these surfaces is provided through the web dashboard described in Section 5.

These maps localize the populations at heightened risk within our subset of severe tropical cyclones. Variation in risk likely reflects the storms sampled and their associated mortality, land composition and geographic features, and the demographic makeup of the blocks affected by storms.

Across storms, the most influential predictors included large block population size, non-Hispanic blocks, male-dominated blocks, predominantly white blocks, and the proportion of males aged 20–34 in a block, summarized by tallying the predictors that ranked consistently high in importance across the boosted-tree models. Socioeconomic covariates contributed little, suggesting limited variation in socioeconomic status across coastal blocks in our sample. These findings ran counter to our predictions and may reflect the ecological study design. See Table 3B for further details.

This study did not examine the individual factors predicting each storm’s mortality and destructive outcomes (Table 3B lists the covariates used to predict mortality per storm). The category-specific tools nonetheless provide fine-resolution risk predictions for the areas most affected by this subset of severe tropical cyclones, and future studies will further elucidate the characteristics of the populations most vulnerable to each storm.

## 4. Discussion

The category-specific risk prediction maps highlight where emergency planning and mitigation are most critical. The results indicate that large block populations, non-Hispanic blocks, blocks with large male populations, and predominantly white blocks carry elevated mortality risk from catastrophic tropical cyclones in the North Atlantic Basin. These findings ran counter to our predictions, likely because census-block analysis cannot capture individual-level variation. The literature shows that racial and ethnic disparities are exacerbated post-storm in terms of adverse health outcomes and increased hospitalizations (Berberian et al., 2022), and that socioeconomic status is a significant predictor of risk from extreme weather events (Zanocco et al., 2022). Evidence on gender-specific risk differences is mixed; determining them requires considering structural inequities, genetics, roles, and behaviors, among other factors (Muñoz- Nieves et al., 2025). Socioeconomic disadvantage also increases vulnerability through more susceptible housing and infrastructure, a lack of transportation for evacuation, and a lack of resources enabling safe evacuation (Bjarnadottir et al., 2011). Our block-level design could not corroborate these findings, as aggregation may have masked individual-level socioeconomic variation. Future studies incorporating the effects of weak infrastructure, resource access, and transportation mechanisms will further strengthen the tool presented in this study.

One existing model that examined hurricane impact similarly demonstrated that strong winds, Asian, African American, and Hispanic populations appeared to be the most vulnerable in terms of economic strife and vulnerability, aligning with some of the findings in this study. Age and socioeconomic status were associated with vulnerability, but the associations were not statistically significant (Burton, 2010). A case study of the fallout from Hurricane Maria in Puerto Rico demonstrated that those with higher SVI (hence greater vulnerability) were more likely to evacuate, and that housing and transportation were significant predictors of evacuation (West, 2023). It would be useful for future investigators to examine social-behavioral mechanisms associated with severe tropical cyclones, as these factors also appear to play a significant role in evacuation tendencies, thereby affecting vulnerability. Other studies have examined population shifts in the composition of coastal and vulnerable communities. While White, mid- to high-income populations appear to be migrating away from coastal communities, their risk remains elevated, alongside poorer, non-White populations that have been moving into coastal areas (Logan & Xu, 2015). This may explain why white populations maintain an elevated risk of mortality, as they may be more likely to avoid evacuation due to a variety of behavioral factors (Logan & Xu, 2015). It would be beneficial to target interventions that assess attitudes and behaviors among communities that have endured numerous hurricane seasons to reduce excess deaths and economic burden.

Additionally, infrastructure is a factor to consider when using this modeling resource to forecast vulnerability. Our findings concur with previous studies that GDP declines ensue post-storm (Frame et al., 2020). This study also emphasizes the importance of analyzing building quality, built environments, floodplain locations, elevation, proximity to water, available resources, trust in government systems, and impervious surfaces, thereby further clarifying coastal vulnerability and informing risk prediction. This was underscored by a 2026 study that outlined a framework for understanding recovery after a storm. They highlight that recovery aid (governmental support), the failure of public infrastructure, labor markets, social assistance, and market dynamics must all be considered to adequately intervene to promote recovery and mitigate community risk (Sou, 2026). Frameworks such as the one proposed by Sou (2026), coupled with the tool generated in this study, can work in tandem to mitigate excess mortality.

Additionally, landscape-specific factors affect the predicted extent of inundation (Anarde et al., 2018). This study shows that it is imperative to account for the dynamics and physical characteristics of the community landscape when assessing where risk is highest. Integrating this risk-prediction tool with infrastructure models offers a holistic view of vulnerability, combining demographic risk with structural exposure to enhance emergency preparedness, mitigation planning, and targeted communication.

Several limitations temper these findings. As an ecological study, this research cannot establish causality or temporality when assessing block-level storm risk, and inference is limited to populations rather than individuals. Our sample of twenty-four of the most severe and costly storms over the past three decades does not represent all storms that have struck the U.S. coast and its territories, and severity categories are unevenly represented; future models should incorporate a larger and more balanced sample of storms. Because analyses are restricted to blocks within 25 miles of the coast, deaths and damage that extended farther inland are not captured. Finally, using 2020 Census data may not reflect the population makeup of communities affected by earlier storms, although archival review of the 1990–2020 Censuses showed the 2020 data to be the most complete and most widely available at the block level across demographic characteristics.

Mortality reported by NOAA is subject to inconsistencies and likely undercounts final death totals and causes of death. Deaths from exacerbated chronic disease, infectious disease spread, or mental distress are often unreported or misclassified on death certificates, so predicted mortality risk is likely conservative. In addition, the population adjustment used to generate block-specific mortality assumes that deaths are distributed proportionally to population across the blocks of a reporting county or state — the finest resolutions NOAA provides.

Economic impacts were reported by NOAA only as aggregate storm totals, so we collected county-level GDP to better gauge the economic burden of storms for blocks; block-level economic inferences are accordingly limited. Because the U.S. Bureau of Labor Statistics provides GDP only for 2001 through 2023, the three storms in our sample occurring before 2001 were excluded from models with the economic outcome, although their aggregate totals remain in the summary statistics.

Both rainfall and inundation data from NOAA required manual, pixel-level digitization in ArcGIS, and block values were computed as means across the pixels composing each block. This averaging may overestimate or underestimate accumulated rainfall at the block level, and predicted inundation is based on our proposed averaging equation rather than observed values.

Where wind swath or rainfall boundaries split a Census block, the block was assigned the hazard category covering more than 50% of its area. For example, a block that was 60% within the hurricane-force wind zone was assigned that value for the entire block. This approach reflects the spatial resolution limits of the processing tools and ensures consistent classification across all blocks.

Finally, the category-level maps average predicted risk across the storms sampled within each category, which masks storm-to-storm variation; storm-specific surfaces are an avenue for future work. Missingness was modest across the individual storm models, ranging from 0.5% to 3%.

## 5. Interactive risk visualization dashboard

To extend the static maps in Figure 2 into a decision-support tool, the category-specific risk surfaces and their underlying block-level covariates have been deployed as a public, interactive web dashboard. The dashboard presents one view per storm category (tropical storm through Category 5). Each view combines three complementary layers rendered by zoom level: a tract-level overview for regional context, a simplified block layer at intermediate scales, and the full-resolution block layer with queryable pop-ups reporting each block’s identifier, predicted mortality risk, and the demographic, socioeconomic, and storm-hazard variables underlying each prediction. All layers are generated directly from the dataset and boosted-tree outputs reported above, ensuring consistency between the manuscript figures and the interactive tool. The dashboard is publicly available at https://hurricanedashboard.com as a companion piece to this article, and the dashboard code and derived map layers are deposited in the MINDS@UW repository (University of Wisconsin–Madison), with a citable DOI to be issued upon completion of repository review.

## 6. Conclusion

This study identifies the populations at greatest mortality risk from extreme tropical cyclones — most notably large-population blocks composed largely of white, non-Hispanic males aged 20-34 years. The boosted tree analysis and a large, comprehensive dataset support population-level inference about which storm factors and demographics contribute to excess mortality risk along the Gulf and East Coasts, Puerto Rico, and the U.S. Virgin Islands, with block-level resolution capturing population composition within the 25-mile buffer at the finest scale available. The resulting risk maps supplement current forecasting and meteorology by identifying where the burden is greatest, and they demonstrate worst-case-scenario risks to guide planning and community-specific mitigation strategies.

These findings open several avenues for further modeling, research, and investigation, particularly for public health and disease surveillance. Storm-specific models should be investigated to determine the effects associated with each storm and how these, in turn, affect mortality. Newly inundated environments, crowded shelters, and destroyed infrastructure precipitate the spread of infectious diseases, including vector-borne, waterborne, and other highly transmissible pathogens. Our findings indicate that excess mortality concentrates in urban coastal communities and varies with storm severity: Puerto Rico, Louisiana, Florida, North Carolina, Virginia, Maryland, New Jersey, and New York consistently show elevated risk, particularly in inlet, peninsula, and sound geographies. Supplemental educational tools such as the risk maps and dashboard developed here can save lives by identifying priority routes for emergency planning, bolstering infrastructure, and optimizing evacuation resources informed by local needs and attitudes. This study provides essential groundwork for modeling the worst-case tropical cyclones projected to impact the U.S. coasts and U.S. territories in the coming years as climate change worsens.

## Data Availability

All data produced in the present study are contained in the manuscript (provided as publicly available urls). Additionally, we have compiled all data in an interactive dashboard, titled 'The Hurricane Dashboard' (https://www.hurricanedashboard.org) where users can download all data there as well.

https://www.ncei.noaa.gov/stormevents/

## Conflict of Interest Statement

The authors declare that they have no conflicts of interest relevant to this manuscript.

## Acknowledgments

The authors thank Mary Ryan Baumann and Silvia Helena Barcellos (School of Medicine and Public Health, University of Wisconsin–Madison), who served on S. Mullins’s thesis committee, for their guidance and feedback. We also thank David Roth (NOAA Weather Prediction Center) and Wallace Hogsett (NOAA National Hurricane Center) for their assistance with archival searches of past storms, and the Department of Population Health Sciences at the University of Wisconsin–Madison for its support. This research received no specific external funding.

## Author contribution

S. Mullins — conceptualization, data curation, formal analysis, visualization, writing – original draft; J. Uelmen — conceptualization, methodology, supervision, formal analysis, writing – review and editing. The georeferencing and supervised pixel-based classification of rainfall and wind-swath imagery were performed by the authors in ArcGIS Pro using its built-in maximum-likelihood and machine-learning classifiers; no generative artificial-intelligence tools were used to generate, analyze, or interpret the study data. Claude (Anthropic) was used to assist with language editing for clarity and readability; all suggested edits were reviewed and approved by the authors, who take full responsibility for the content of this manuscript. This research received no specific external funding.

## Open Research

### Data Availability Statement

All primary data used in this study are publicly available. Human mortality data are available from the National Weather Service’s (NWS) Storm Events Database. Demographic and socioeconomic data are from the U.S. Census Bureau 2020 Decennial Census (Demographic and Housing Characteristics file) and American Community Survey (U.S. Census Bureau, 2020, 2023). Tropical-cyclone landfall characteristics and wind-swath and rainfall products are from the NOAA National Hurricane Center Tropical Cyclone Reports and Data Archive (National Hurricane Center, 2026; NHC Data Archive, 2026); cost estimates are from the NOAA National Centers for Environmental Information (National Centers for Environmental Information, 2025); weather-related mortality is from the NOAA/NWS Storm Events Database. The Social Vulnerability Index is from the CDC/ATSDR (CDC, 2024). County gross domestic product is from the U.S. Bureau of Labor Statistics. The derived, block-level analysis dataset and all processing and modeling scripts (R and JMP) that generated the tables and figures, together with the code underlying the interactive dashboard, are archived at MINDS@UW repository [DOI/URL to be inserted at acceptance], under a [CC-BY 4.0 / open] license. The interactive dashboard (Section 5) is available at https://hurricanedashboard.com and is deposited in the MINDS@UW repository (University of Wisconsin–Madison); its DOI is pending repository review [dashboard DOI to be inserted]. Analyses were performed in R (R Core Team), JMP Student Edition 18.2.1 (SAS Institute), and ArcGIS Pro (Esri); each is cited in the reference list.

## 8. Supplemental Materials

**Table 1A.**
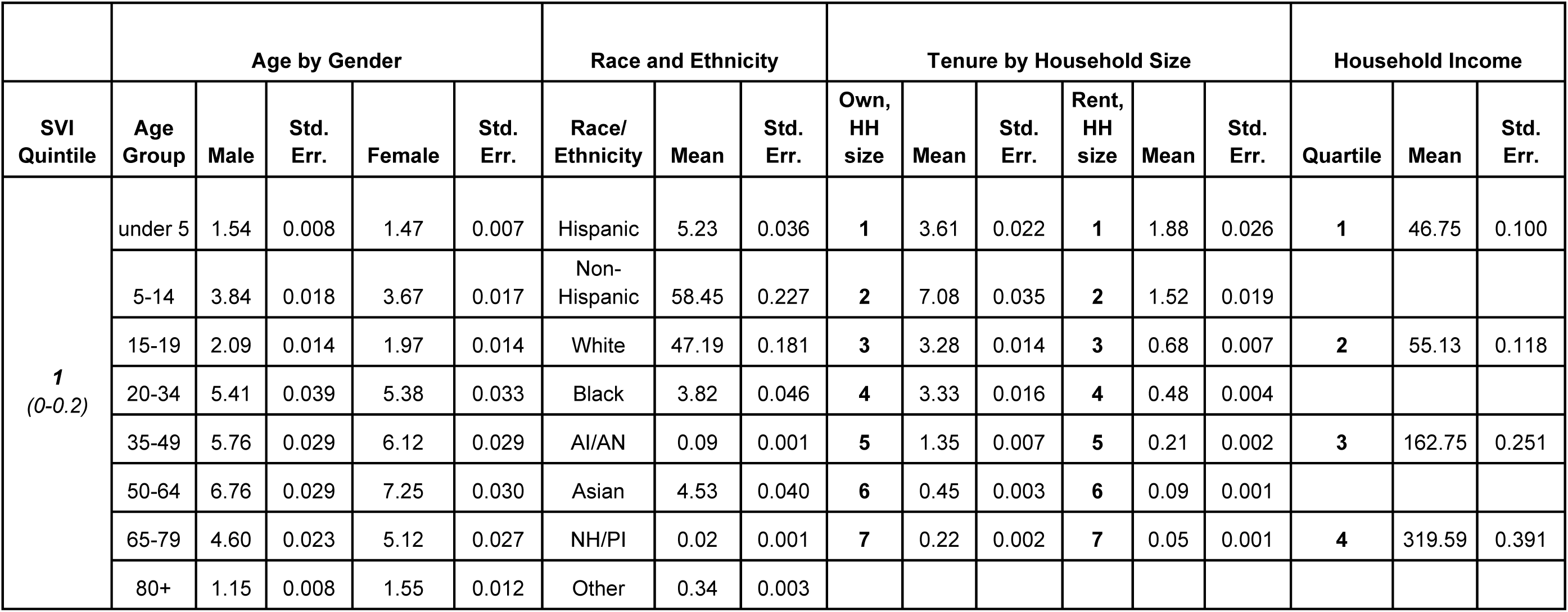
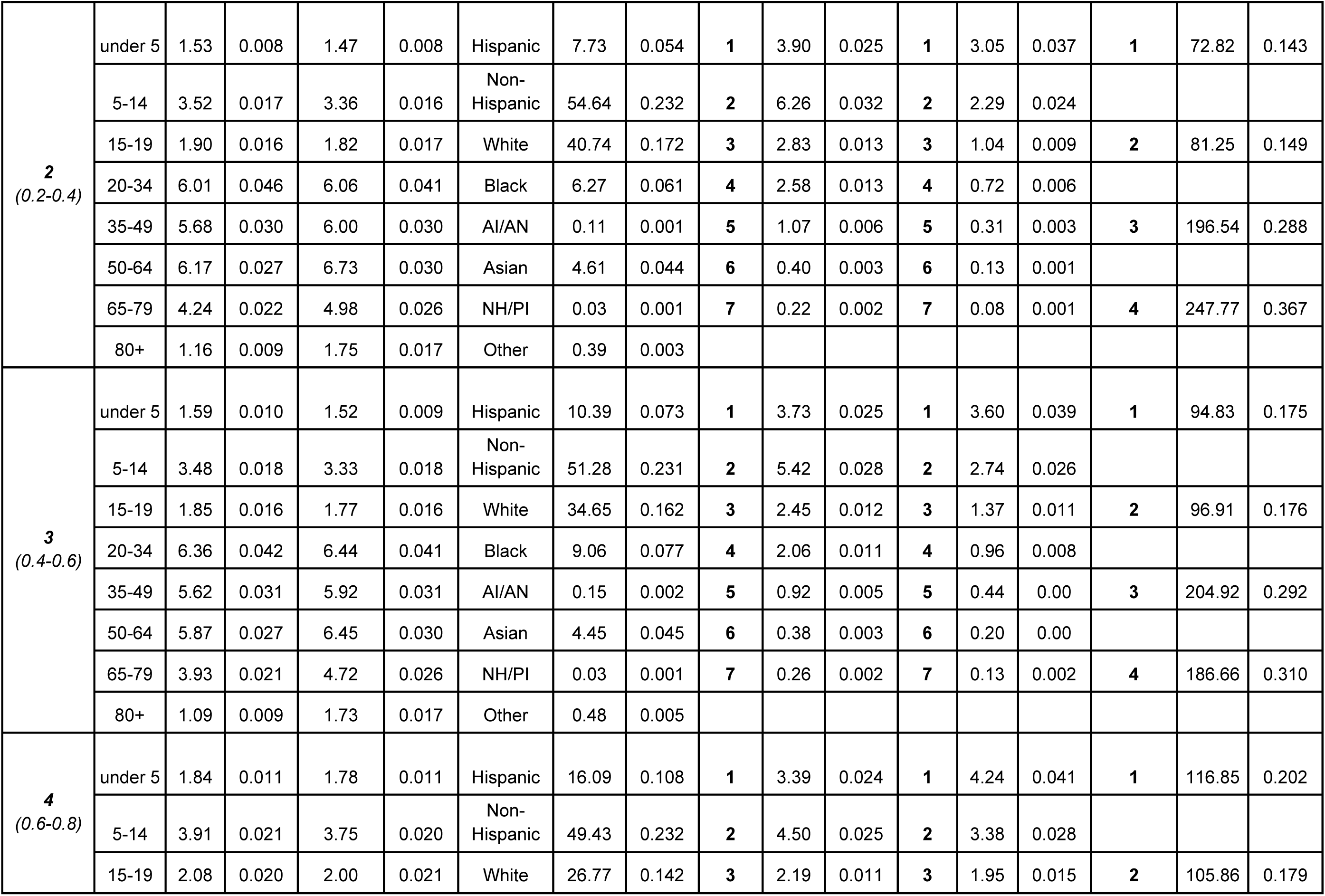
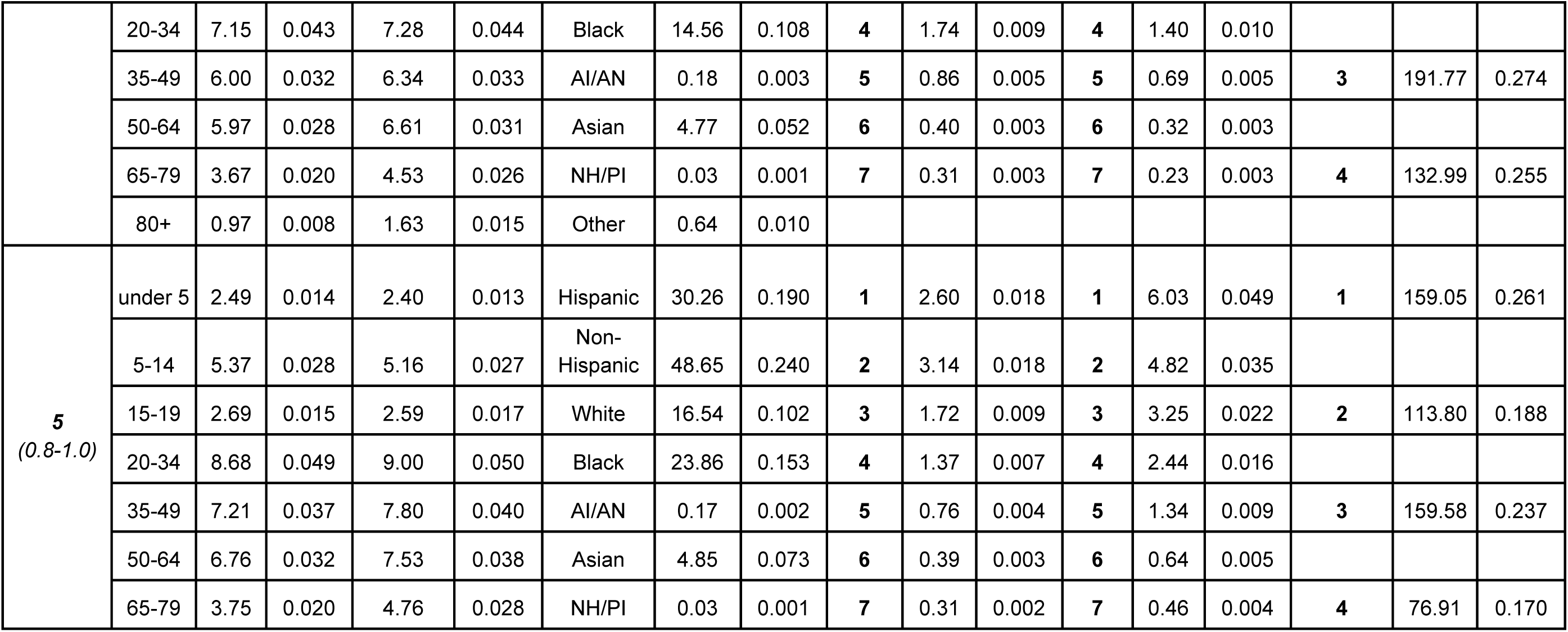
Summary statistics of the entire dataset, stratified by SVI quintiles. Does not include Puerto Rico, the U.S. Virgin Islands, or Connecticut due to insufficient SVI reporting. See Table 4B for breakdown by state, including Puerto Rico, and supplemental Table 2B for U.S. Virgin Islands breakdown due to differences in reporting by the U.S. Census Bureau. Means and standard errors are reported for each characteristic, where AI/AN is American Indian or Alaskan Native, NH/PI is Native Hawaiian or Pacific Islander, and HH size signifies household size.

**Table 1B:** Summary of data sources utilized and associated acronyms and contents.

| **Data Source** | **Contents** |
| --- | --- |
| National Oceanic and Atmospheric Administration (NOAA) | All extreme weather data and hurricane summary reports |
| National Hurricane Center (NHC) | Inundation data, archival hurricane data and associated characteristics |
| National Center for Environmental Information (NCEI @ NOAA) | https://www.ncei.noaa.gov/access/billions/dcmi.pdf (mortality findings) |
| - Inundation | https://www.nhc.noaa.gov/nhcexit.php?outurl=http://journals.ametsoc.org/doi/abs/10.1175/WCAS-D-14-00049.1 |
| - Wind swaths and best tracks | https://www.nhc.noaa.gov/data/ |
| U.S. Census Bureau | All socioeconomic and demographic data |
| - American Community Survey | Socioeconomic household data |
| - Demographic and Housing Characteristics Survey | Demographics and socioeconomic predictor variables |
| Social Vulnerability Index (SVI) | 2022 CDC compiled dataset that measures population vulnerability at census-tract level based on specific economic and demographics |

**Table 2A.**
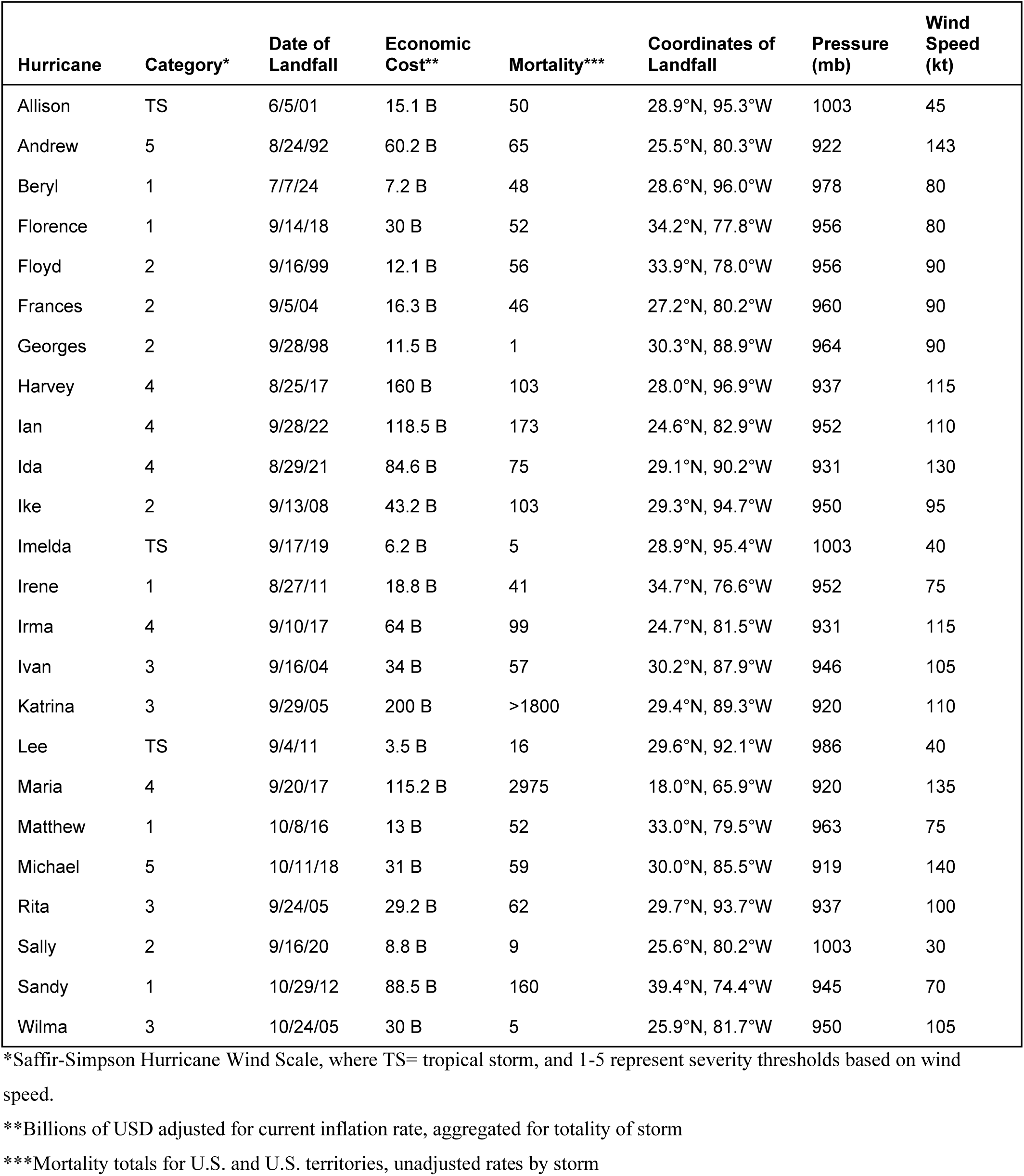
Summary data of landfall characteristics associated with each tropical cyclone in our sample. All data were compiled from NOAA summary files published after each storm.

**Table 2B.** U.S. Virgin Islands Summary Statistics. U.S. Census Bureau reporting is inconsistent with reporting for the contiguous U.S. and Puerto Rico. Owning or renting is stratified by race, median age is available by gender, totals for owning and renting, and average household size are available in accordance with the 2020 census. Means and standard errors are reported for each characteristic.

| Tenure by Race |  |  |  | Median Age by Gender |  |  | Tenure |  |  | Avg. Household Size |  |
| --- | --- | --- | --- | --- | --- | --- | --- | --- | --- | --- | --- |
|  |  | Mean | Std. Err. |  | Mean | Std. Err. |  | Mean | Std. Err. | Mean | Std. Err. |
| Own | Black | 3.086 | 0.1147 | Male | 24.49 | 0.465 | Own | 5.52 | 0.236 | 2.47 | 0.034 |
|  | White | 0.877 | 0.0374 | Female | 24.69 | 0.468 | Rent | 6.18 | 0.363 |  |  |
|  | AI/AN | 0.016 | 0.0020 |  |  |  |  |  |  |  |  |
|  | Asian | 0.053 | 0.0053 |  |  |  |  |  |  |  |  |
|  | NH/PI | 0.001 | 0.0004 |  |  |  |  |  |  |  |  |
|  | Other | 0.173 | 0.0104 |  |  |  |  |  |  |  |  |
| Rent | Black | 3.456 | 0.1808 |  |  |  |  |  |  |  |  |
|  | White | 0.836 | 0.0396 |  |  |  |  |  |  |  |  |
|  | AI/AN | 0.021 | 0.0023 |  |  |  |  |  |  |  |  |
|  | Asian | 0.056 | 0.0066 |  |  |  |  |  |  |  |  |
|  | NH/PI | 0.001 | 0.0004 |  |  |  |  |  |  |  |  |
|  | Other | 0.201 | 0.0131 |  |  |  |  |  |  |  |  |

**Table 3A.** Inclusion and exclusion summaries and justification of excess mortality associated with tropical cyclones reported by NOAA for the U.S. and U.S. territories.

| Hurricane | Death Total | Mortality Included* | Mortality Excluded** |
| --- | --- | --- | --- |
| Beryl | 48 | 48 | 0 |
| Ian*** | 173 | 162 | 11 |
| Ida*** | 75 | 60 | 15 |
| Sally | 9 | 9 | 0 |
| Imelda | 5 | 0 | 5 |
| Michael | 59 | 44.25 | 14.75 |
| Florence | 52 | 41 | 11 |
| Maria**** | 2975 | 2975 | 0 |
| Irma | 99 | 96 | 3 |
| Harvey | 103 | 96 | 7 |
| Matthew | 52 | 26 | 26 |
| Sandy | 160 | 155.6 | 4.4 |
| Lee | 16 | 3 | 13 |
| Irene | 41 | 21 | 20 |
| Ike | 112 | 84 | 28 |
| Wilma | 5 | 5 | 0 |
| Katrina**** | 1800 | 1085 | 307 |
| Rita | 62 | 55 | 7 |
| Ivan | 57 | 34.2 | 22.8 |
| Frances | 46 | 45 | 1 |
| Allison | 50 | 23 | 27 |
| Floyd | 56 | 55 | 1 |
| Georges | 1 | 1 | 0 |
| Andrew | 65 | 55 | 6 |

\*Mortality included in buffer zone of interest and with sufficient data on location reported by NOAA
\*\*Mortality excluded because fatalities were located outside of the 25-mile buffer, or there was insufficient data on location
\*\*\*Deaths are averaged due to extrapolation of spatial reporting by NOAA
\*\*\*\*Approximate deaths, likely underreported

**Table 3B.**
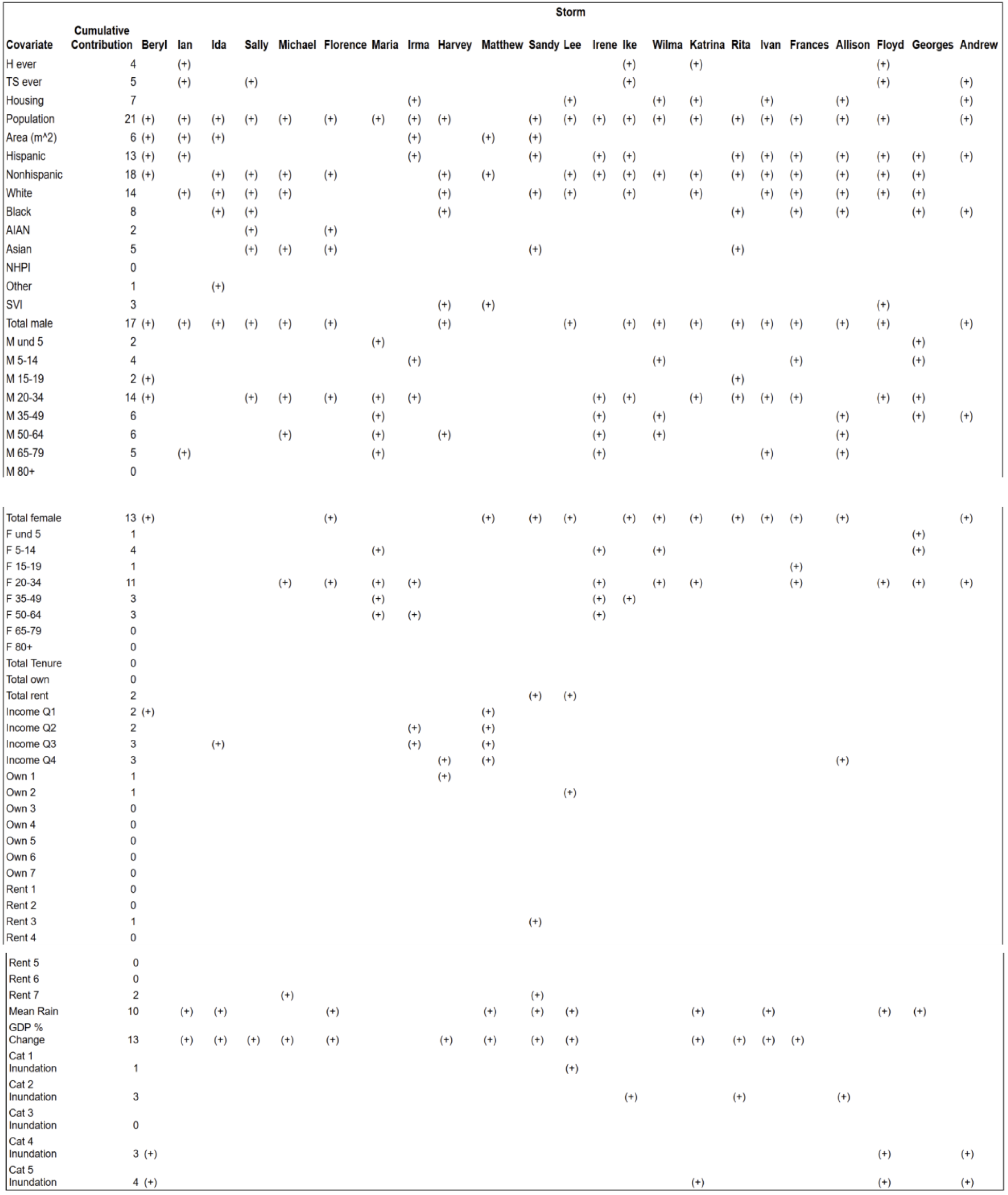
Storm-specific covariate grid showing the cumulative contribution of each covariate to predicting mortality risk, summed across all storms analyzed in the sample. Block population total, gender, ethnicity, and predominantly white blocks appear to be correlated with mortality the most across storms.

**Table 4A.** Storm-specific boosted tree results, including R^2^ and root average square error (RASE). These results inform the category-specific tools and associated maps. Across individual storms, R^2^ demonstrates relatively good model fitness and generally low RASE suggests relatively accurate predictive power.

| **Storm** | **$R^2$** | **RASE** |
| --- | --- | --- |
| **Wilma** | 0.999 | 0.0000018 |
| **Maria** | 0.994 | 0.0028948 |
| **Irene** | 0.992 | 0.0000017 |
| **Ike** | 0.948 | 0.0004675 |
| **Sandy** | 0.932 | 0.0001024 |
| **Ivan** | 0.931 | 0.0003925 |
| **Irma** | 0.928 | 0.0000699 |
| **Florence** | 0.923 | 0.0003579 |
| **Rita** | 0.919 | 0.0004504 |
| **Katrina** | 0.915 | 0.0072647 |
| **Allison** | 0.879 | 0.0012738 |
| **Lee** | 0.870 | 0.0000559 |
| **Floyd** | 0.866 | 0.0002218 |
| **Andrew** | 0.843 | 0.0004433 |
| **Frances** | 0.804 | 0.0004249 |
| **Sally** | 0.710 | 0.0001759 |
| **Harvey** | 0.671 | 0.0024359 |
| **Beryl** | 0.631 | 0.0012207 |
| **Georges** | 0.613 | 0.0000310 |
| **Ida** | 0.369 | 0.0015790 |
| **Michael** | 0.333 | 0.0069659 |
| **Matthew** | 0.250 | 0.0099573 |
| **Ian** | 0.229 | 0.0079945 |

**Table 4B.**
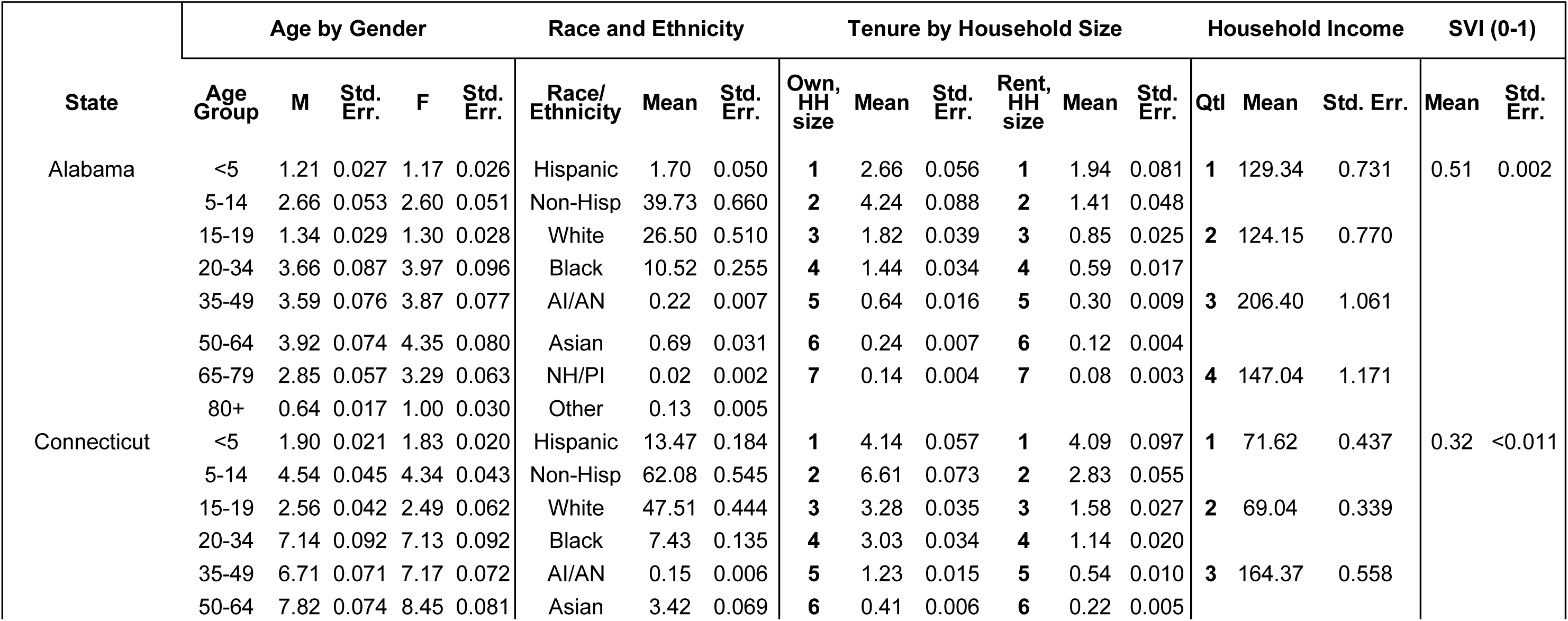
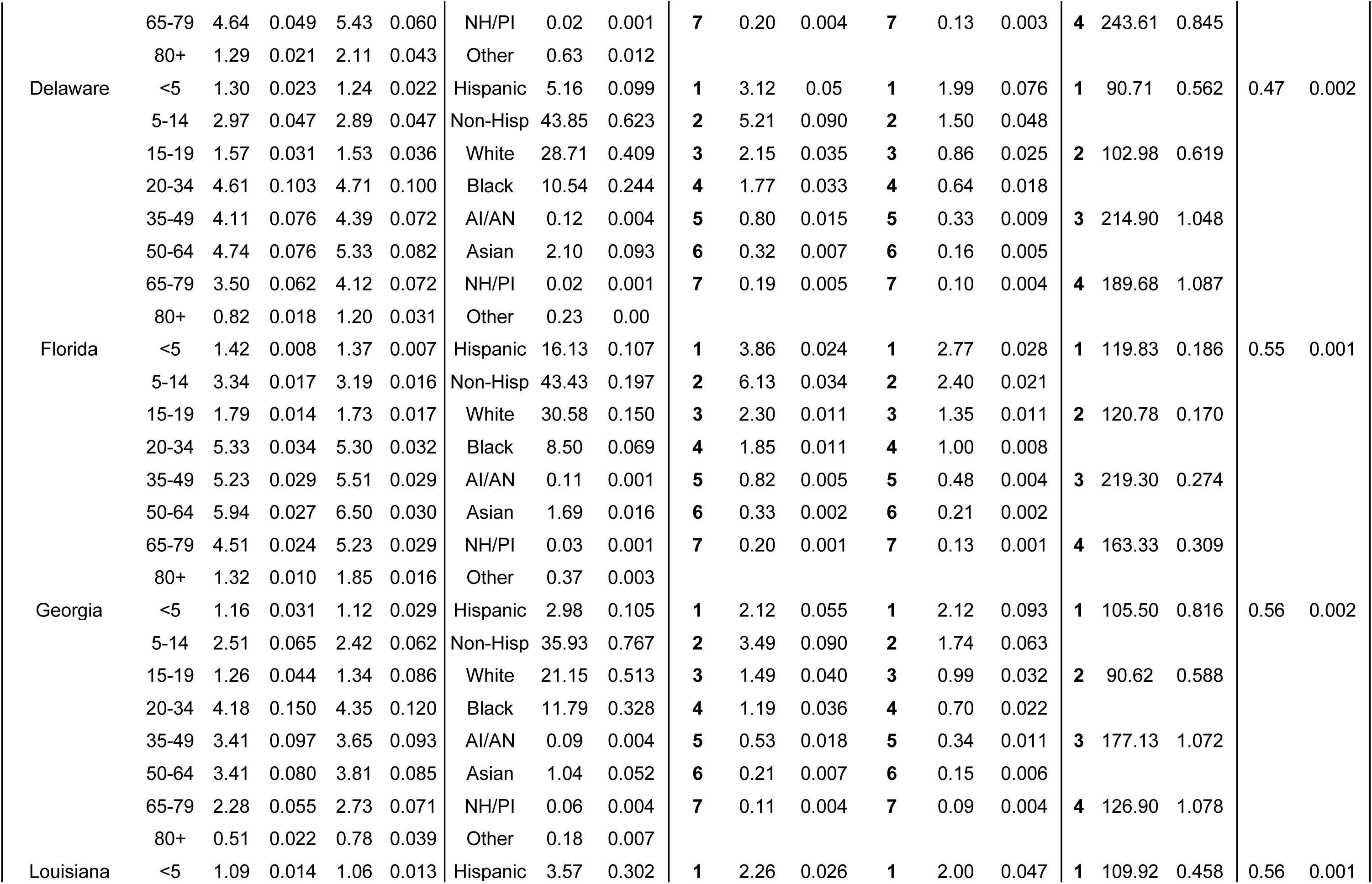
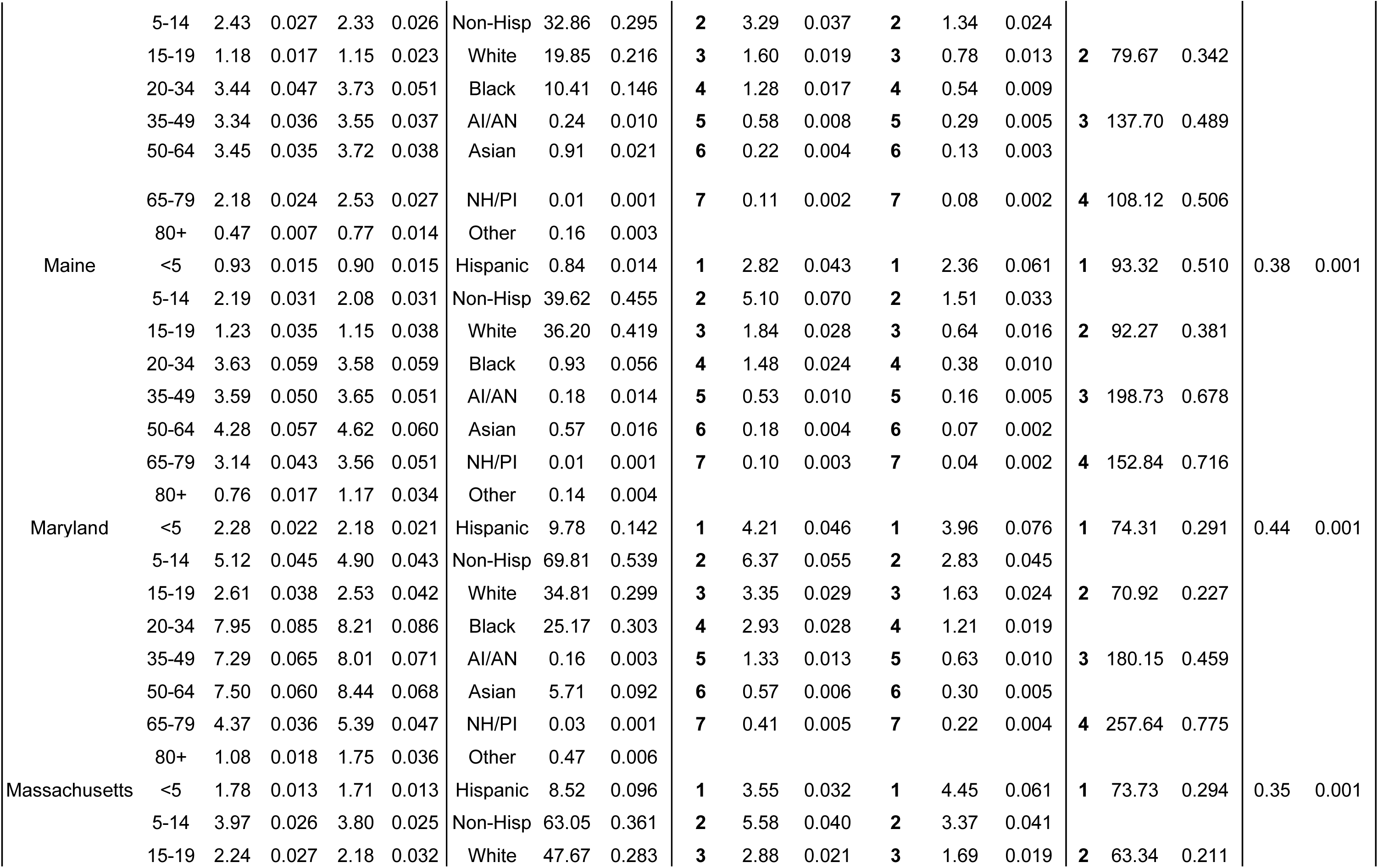
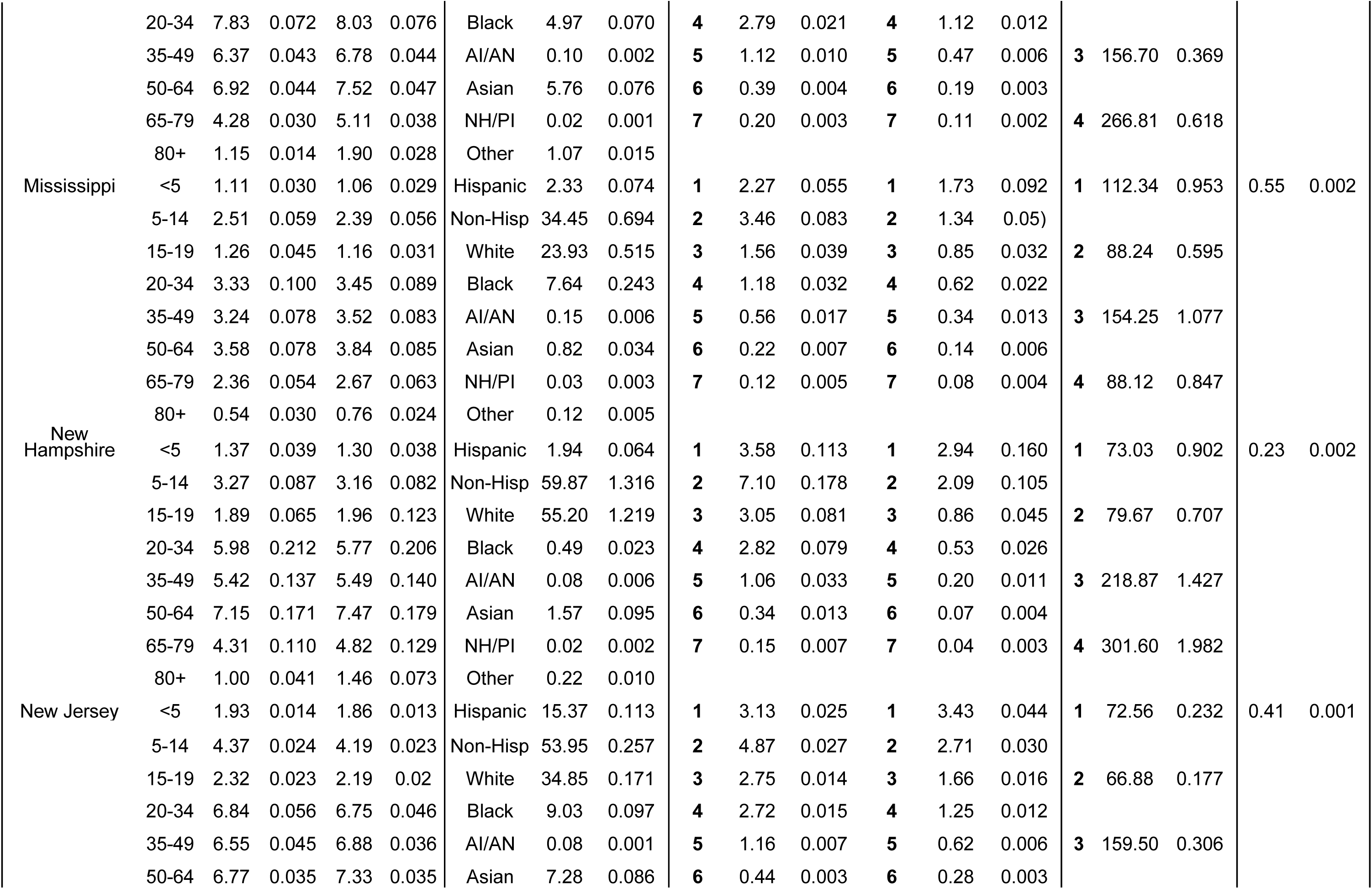
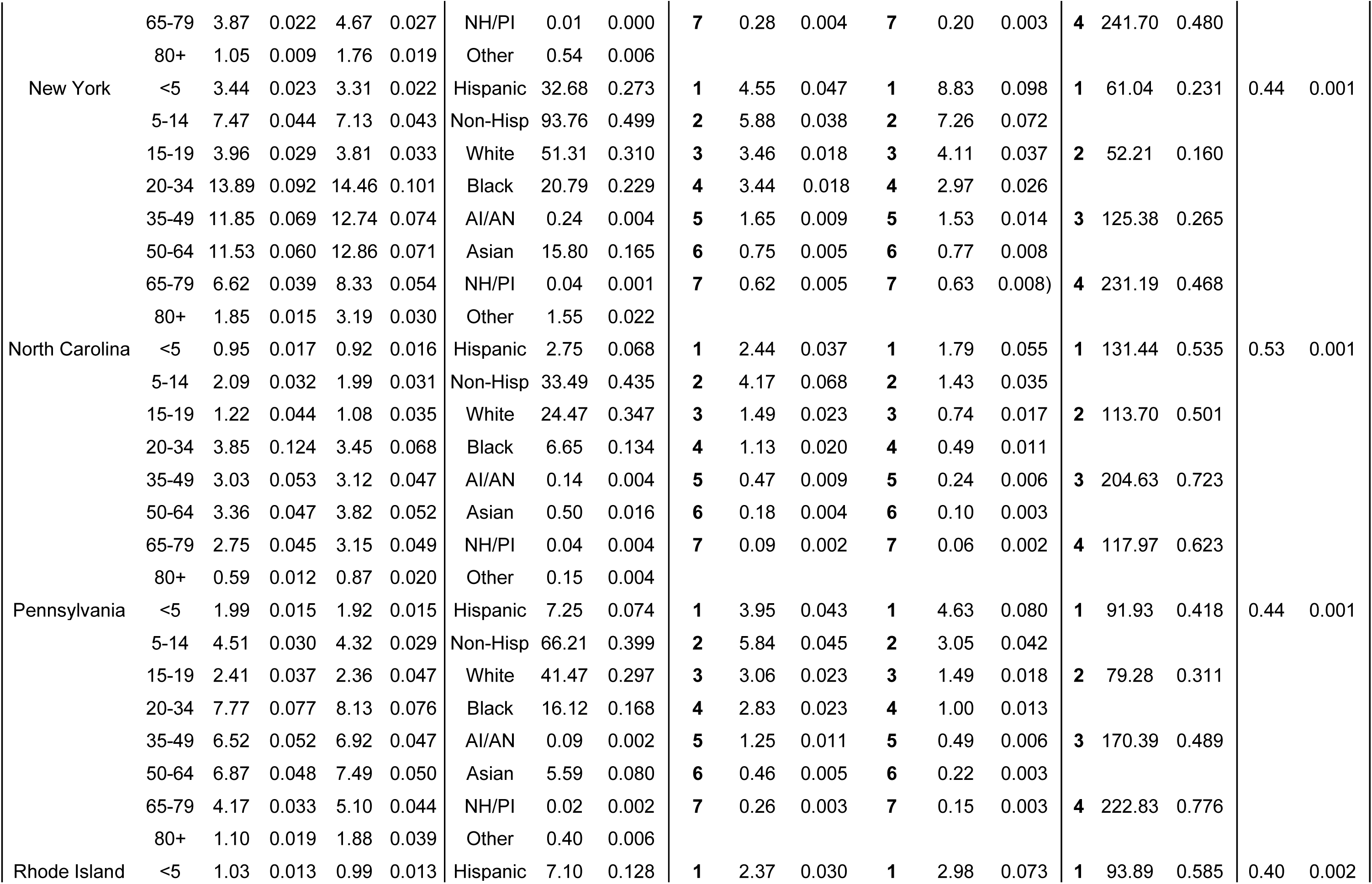
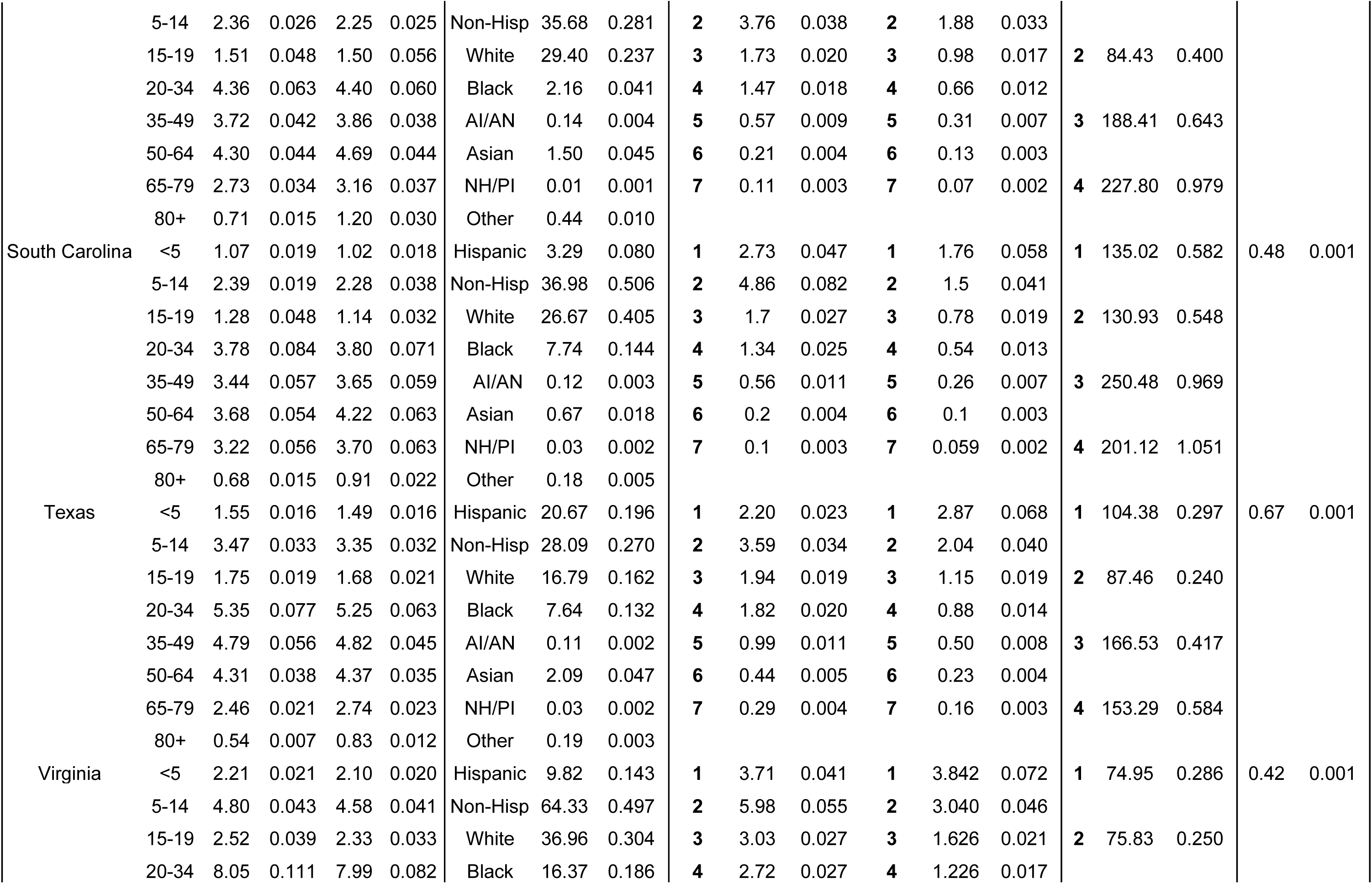
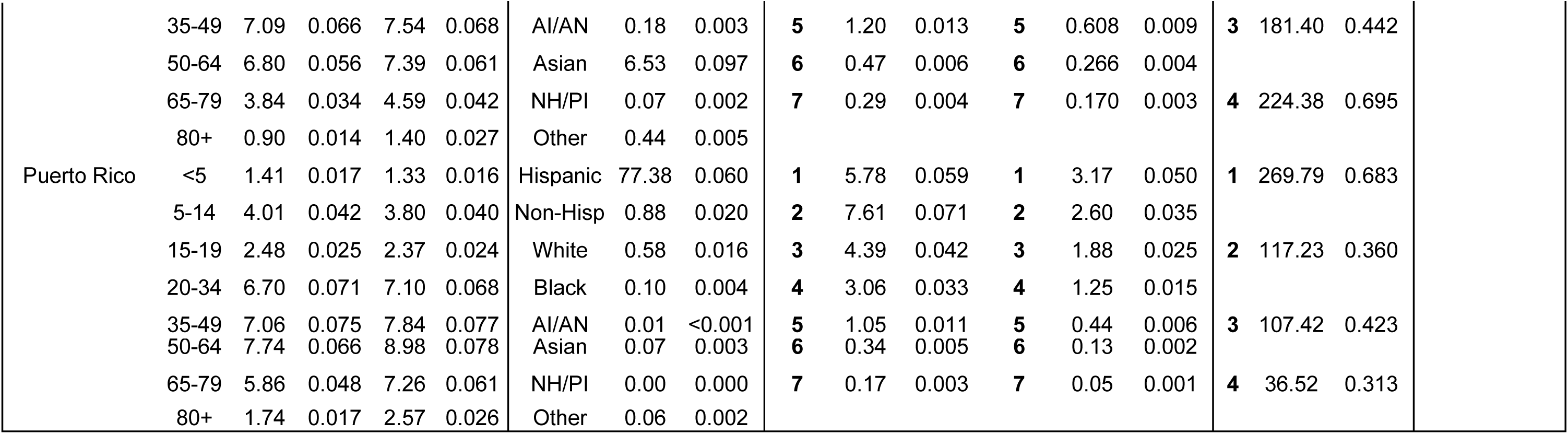
Summary Statistics by State, including Puerto Rico. See the U.S. Virgin Islands summary in Table 2B due to differences in reporting by the U.S. Census Bureau. Means and standard errors are reported for each characteristic, where Non-Hisp is Non- Hispanic, AI/AN is American Indian or Alaskan Native, NH/PI is Native Hawaiian or Pacific Islander, and HH size signifies household size. M represents male averages, and F represents female averages for the Age by Gender category. Qtl. stands for income quartile.

## Supplemental limitations in totality

We acknowledge that this tool has limitations. As an ecological study, this research cannot establish causality or establish temporality when assessing block-level risk associated with storms. Furthermore, while this study is incredibly robust and has sufficient statistical power, statistical significance is difficult to elucidate and the effects of our covariates are likely underestimated. Sampling twenty-four of the most severe and costly storms over the past thirty years is not comprehensive of all storms that have ravaged the U.S. coast /U.S. territories.

Furthermore, there were unequal numbers of sampled storms within each category of the Saffir- Simpson scale. This limits our ability to accurately and equally compare storm burden, specifically among Tropical Storms and Category 5 cyclones, which only contributed a total of five of the twenty-four storms. Future models should incorporate a larger sample of storms, varying in categorization severity. As mentioned in our methods, utilizing 2020 Census data may not correspond with the population makeup of communities affected by earlier storms. However, through archival searches of previous Censuses spanning 1990-2020, we found that 2020 data was the most complete and interpretable, and most widely available at the block-level resolution across demographic characteristics. Additionally, this study samples aggregate data given the expansive geographic regions that were covered and the nature of ecological and weather- dependent data. As such, we cannot draw inferences on individual-level vulnerabilities and risk. To address this, we filtered population demographics at the block level, which is the finest resolution available from the U.S. Census Bureau.

Mortality data reported by NOAA generates a vast array of inconsistencies and likely underreporting of final death totals and causes of death. This study does not account for any mortality that ensued due to exacerbation of chronic disease, infectious disease spread, or mental distress related deaths, due to a lack of reporting or misclassification of deaths on death certificates, for example. As such, mortality is likely underestimated in the study and the final rates on mortality are greater than what this study can conclude. Additionally, the population adjustment that we performed to generate a block-specific mortality rate assumes that each block within a state or county (since these are the two resolutions that were provided by NOAA) is equally affected by the mortality rate. To address this, we adjusted for the specific population in a given block. Despite this, the assumption that mortality rate is equal among blocks is likely untrue and further data should be collected in future studies to ascertain risk at fine resolutions.

Economic data for each storm was aggregated and reported by NOAA as a total for the storm. This is not informative for estimating block-level risk, so we also collected county-level GDP data in order to better gauge the economic burden of storms for blocks. We cannot make assumptions about the economic toll at the block level due to the nature of county-level reporting by the U.S. Bureau of Labor Statistics. Additionally, this data source only contained GDP data spanning 2001 through 2023. The storms in our sample occurring prior to 2001, totaling three storms, were excluded from the analysis with the economic outcome. However, the aggregate totals are still included in summary statistics.

There were inconsistencies in wind swath and rainfall reporting by NOAA for storms occurring after 2009 versus prior to 2009. NOAA’s generated wind swath files for the more recent storms included delineations of wind speed at three different knot levels (34, 50, and 64 knots) whereas any cyclones that made landfall prior to 2009 were only delineated between “hurricane force winds” and “tropical storm force winds.” Due to the differences in reporting, we standardized wind data to either “hurricane force winds” or “tropical storm force winds”, which aggregates the heterogeneity of 34 and 50 knot force winds. We made comparisons then among these two categories as opposed to three. Both rainfall and inundation data from NOAA necessitated manual digitization at the pixel level in ArcGIS software, where blocks are composed of various pixels. To gain an understanding of the rainfall and predicted inundation that accumulated in each block for each storm, we calculated mean rainfall totals per block based on the totality of the pixel values corresponding to rainfall totals. This may have resulted in underestimations as well as over estimations in the rainfall that accumulated at the block level, which may skew the risk associated with that block. For predicted inundation, we proposed an equation to gain a better estimate at the block level, yet this is still based on proposed averages and not necessarily true values.

Finally, when overlaying the census block buffer with the wind swaths and rainfall map, at times the maps split through blocks. In cases where storm-impact boundaries did not align cleanly with Census block boundaries, we assigned values based on which hazard category covered more than 50 percent of the block. For example, if the line between hurricane-force and tropical-storm-force winds split a block 40 percent versus 60 percent, the block was classified according to the category covering the larger share. Thus, a block that was 60 percent within the hurricane-force wind zone was assigned that value for the entire block. This approach reflects the spatial resolution limits of ArcGIS processing tools and ensures consistent classification across all blocks.

## Notes

### Competing Interest Statement

The authors have declared no competing interest.

### Author Declarations

The source human mortality data is publicly available from the National Centers for Environmental Information via this URL: https://www.ncei.noaa.gov/stormevents/.

## References

Alexander, M. A. (2020). The Response of the Northwest Atlantic Ocean to Climate Change. Journal of Climate Volume 33(*2*), 405–428. 10.1175/JCLI-D-19-0117.1

Anarde, K. A., Kameshwar, S., Irza, J. N., Nittrouer, J. A., Lorenzo-Trueba, J., Padgett, J. E., Sebastian, A., & Bedient, P. B. (2018). Impacts of Hurricane Storm Surge on Infrastructure Vulnerability for an Evolving Coastal Landscape. Natural Hazards Review, 19(1), 04017020. 10.1061/(ASCE)NH.1527-6996.0000265

Berberian, A.G., Gonzalez, D.J.X. & Cushing, L.J. (2022). Racial Disparities in Climate Change-Related Health Effects in the United States. Curr Envir Health Rpt 9, 451–464.10.1007/s40572-022-00360-w

Beven, J. L. II, Berg, R., & Hagen, A. (2019). Tropical Cyclone Report: Hurricane Michael(AL142018), 7-11 October 2018. National Hurricane Center. https://www.nhc.noaa.gov/data/tcr/AL142018_Michael.pdf

Bjarnadottir, S., Li, Y., & Stewart, M. G. (2011). Social vulnerability index for coastal communities at risk to hurricane hazard and a changing climate. Natural Hazards, 59(2), 1055–1075. 10.1007/s11069-011-9817-5

Burton, C. G. (2010). Social Vulnerability and Hurricane Impact Modeling. Natural Hazards Review, 11(2), 58–68. 10.1061/(ASCE)1527-6988(2010)11:2(58)

CDC. (2024, October 22). Social Vulnerability Index. Place and Health - Geospatial Research, Analysis, and Services Program (GRASP). https://www.atsdr.cdc.gov/place-health/php/svi/index.html

Cowan, K. N., Zavala, D. E., Suarez, E., Lopez-Rodriguez, J. A., & Alvarez, O. (2025). Excess mortality and associated community risk factors related to hurricane Maria in Puerto Rico. Environmental Research: Health, 3(1), 015014. 10.1088/2752-5309/adac03

Delworth, T. L., Zeng, Fanrong, Vecchi, Gabriel A., Yang, Xiaosong, Zhang Liping, & Zhang, Rong (2016). The North Atlantic Oscillation as a driver of rapid climate change in the Northern Hemisphere. Nature Geoscience. (9), 509–512. https://www.nature.com/articles/ngeo2738

Dosa, D., Feng, Z., Hyer, K., Brown, L. M., Thomas, K., & Mor, V. (2010). Effects of Hurricane Katrina on Nursing Facility Resident Mortality, Hospitalization, and Functional Decline. Disaster Medicine and Public Health Preparedness, 4(S1), S28–S32. 10.1001/dmp.2010.11

Frame, D. J., Wehner, M. F., Noy, I., & Rosier, S. M. (2020). The economic costs of Hurricane Harvey attributable to climate change. Climatic Change, 160(2), 271–281. 10.1007/s10584-020-02692-8

Franco, B. C., Defeo, Omar, Moller, & Osmar O. (2020). Climate change impacts on the atmospheric circulation, ocean, and fisheries in the southwest South Atlantic Ocean: A review. Climatic Change. 162, 2359–2377. 10.1007/s10584-020-02783-6

Hori, M., & Schafer, M. J. (2009). Social costs of displacement in Louisiana after Hurricanes Katrina and Rita. Population and Environment. 31. 64–86. 10.1007/s11111-009-0094-0

Liang, S. Y., & Messenger, N. (2018). Infectious diseases after hydrologic disasters. Emergency Medicine Clinics of North America, 36(4), 835–851. 10.1016/j.emc.2018.07.002

Logan, J. R., & Xu, Z. (2015). Vulnerability to Hurricane Damage on the U.S. Gulf Coast Since 1950. Geographical Review, 105(2), 133–155. 10.1111/j.1931-0846.2014.12064.x

McKinney, N., Houser, C., & Meyer-Arendt, K. (2011). Direct and indirect mortality in Florida during the 2004 hurricane season. International Journal of Biometeorology, 55(4), 533–546. 10.1007/s00484-010-0370-9

Muñoz-Nieves, C., Greaves, L., Huber, E., Brabete, A. C., Wolfson, L., & Poole, N. (2025). Sex and Gender Influences on the Impacts of Disasters: A Rapid Review of Evidence. International Journal of Environmental Research and Public Health, 22(9), 1417. 10.3390/ijerph22091417

Nam, C., Cha, T., Casas E., Klotzbach P., Young K., Silvers L., Bell M. (2026). CSU Tropical Cyclone Impact Probabilities. CSU Tropical Cyclones, Radar, Atmospheric Modeling, and Software Team. https://tropical.colostate.edu/TC_impact.html

National Centers for Environmental Information. (2025). Costliest U.S. tropical cyclones (NOAA Technical Report). National Oceanic and Atmospheric Administration. https://www.ncei.noaa.gov/access/billions/dcmi.pdf

National Hurricane Center. (2026). Tropical cyclone reports archive. National Oceanic and Atmospheric Administration. https://www.nhc.noaa.gov/data/tcr/index.php

NHC Data Archive. (2026). Retrieved May 29, 2025, from https://www.nhc.noaa.gov/data/

Salim, M. Z., Qiang, Y., Dixon, B., & Collins, J. (2024). A Disparate Disaster: Spatial Patterns of Building Damage Caused by Hurricane Ian and Associated Socio-Economic Factors. Remote Sensing, 16(20), Article 20. 10.3390/rs16203792

Schwartz, R. M., Liu, B., Lieberman-Cribbin, W., & Taioli, E. (2017). Displacement and mental health after natural disasters. The Lancet Planetary Health, 1(8), e314. 10.1016/S2542-5196(17)30138-9

Seidel, D., Wurster, S., Jenks, J. D., Sati, H., Gangneux, J.-P., Egger, M., Alastruey-Izquierdo, A., Ford, N. P., Chowdhary, A., Sprute, R., Cornely, O., Thompson, G. R., Hoenigl, M., & Kontoyiannis, D. P. (2024). Impact of climate change and natural disasters on fungal infections. The Lancet Microbe, 5(6), e594–e605. 10.1016/S2666-5247(24)00039-9

Shultz, J. M., Kossin, J. P., Ettman, C., Kinney, P. L., & Galea, S. (2018). The 2017 perfect storm season, climate change, and environmental injustice. The Lancet Planetary Health, 2(9), e370–e371. 10.1016/S2542-5196(18)30168-2

Smith, T. J., Anderson, G. H., Balentine, K., Tiling, G., Ward, G. A., & Whelan, K. R. T. (2009). Cumulative impacts of hurricanes on Florida mangrove ecosystems: Sediment deposition, storm surges and vegetation. Wetlands, 29(1), 24–34. 10.1672/08-40.1

Solís, D., Thomas, M., & Letson, D. (2010). An empirical evaluation of the determinants of household hurricane evacuation choice. Journal of Development and Agricultural Economics, 2(*3*), 188–196.

Sou, G. (2026). A place-based framework to understand family disaster recovery. International Journal of Disaster Risk Reduction, 132, 105962. 10.1016/j.ijdrr.2025.105962

U.S. Census Bureau. (2010). Average household size of occupied housing units by tenure (DEC Summary File 1, Table H12). Retrieved January 16, 2025, from https://data.census.gov/table/DECENNIALSF12010.H12

U.S. Census Bureau. (2020). Hispanic or Latino, and not Hispanic or Latino by race (DEC 118th Congressional District Summary File, Table P9). Retrieved January 16, 2025, from https://data.census.gov/table/DECENNIALCD1182020.P9

U.S. Census Bureau. (2023). Age and sex (American Community Survey, ACS 1-Year Estimates, Table S0101). Retrieved January 16, 2025, from https://data.census.gov/table/ACSST1Y2023.S0101

U.S. Census Bureau. (2023). Demographic characteristics for occupied housing units (American Community Survey, ACS 1-Year Estimates, Table S2502). Retrieved January 16, 2025, from https://data.census.gov/table/ACSST1Y2023.S2502

U.S. Census Bureau. (2023). Income in the past 12 months (in 2023 inflation-adjusted dollars) (American Community Survey, ACS 1-Year Estimates, Table S1901). Retrieved January 16, 2025, from https://data.census.gov/table/ACSST1Y2023.S1901

Watson, J. T., Gayer, M., & Connolly, M. A. (2007). Epidemics after Natural Disasters. Emerging Infectious Diseases, 13(1), 1–5. 10.3201/eid1301.060779

West, J. (2023). Social vulnerability and population loss in Puerto Rico after Hurricane Maria. Population and Environment, 45(2), 8. 10.1007/s11111-023-00418-3

Zane, D. F., Bayleyegn, T. M., Hellsten, J., Beal, R., Beasley, C., Haywood, T., Wiltz-Beckham, D., & Wolkin, A. F. (2011). Tracking Deaths Related to Hurricane Ike, Texas, 2008. DISASTER MEDICINE AND PUBLIC HEALTH PREPAREDNESS, 5(1), 23–28. 10.1001/dmp.2011.8

Zanocco C., Flora J., Boudet H. (2022). Disparities in self-reported extreme weather impacts by race, ethnicity, and income in the United States. PLOS Climate 1(6): e0000026. 10.1371/journal.pclm.0000026

Zhang, F., Orton, P. M., Madajewicz, M., Jagupilla, S. C. K., & Bakhtyar, R. (2020). Mortality during Hurricane Sandy: The effects of waterfront flood protection on Staten Island, New York. Natural Hazards, 103(1), 57–85. 10.1007/s11069-020-03959-0

